# Bedtime Brain State Predicts the Impact of Closed-Loop Auditory Stimulation on Sleep and Cognition

**DOI:** 10.1101/2025.11.12.25340010

**Authors:** Yashar Kiarashi, Jonathan Berent, Mohsen Motie-Shirazi, Qiao Li, Andrew X. Stewart, Prabhjyot Saini, Daniela Grimaldi, Kathryn J. Reid, Phyllis C. Zee, Ken A. Paller, Allan I. Levey, Gari D. Clifford

## Abstract

**Study Objectives:** Sleep interventions targeting slow-wave activity (SWA) show heterogeneous effects across individuals. We investigated whether pre-sleep brain states predict responsiveness to auditory stimulation and associated cognitive benefits.

**Methods:** Twenty-eight healthy adults (19-40 years) completed three overnight laboratory visits under mild sleep restriction. Pre-sleep EEG recordings captured spectral band power before each night. Participants received auditory stimulation or sham on two randomized nights with polysomnography monitoring. Memory recall and sustained attention were assessed at multiple timepoints the following day. Machine learning models using normalized EEG features and transfer learning with pre-trained sleep architectures (SleepNet, DeepSleepNet, TinySleepNet) predicted responsiveness.

**Results:** Auditory stimulation significantly enhanced SWA (*p* < 0.01) in over 90% of participants. Pre-sleep alpha and theta power correlated with SWA enhancement (alpha: *r* = 0.436; theta: *r* = 0.329; both *p* < 0.01), as did sleep onset latency (*r* = 0.429, *p* < 0.01). Temporal embeddings derived from 8 minutes of pre-sleep EEG before lights off predicted individual responsiveness to auditory stimulation with 80% accuracy (AUC = 0.93).

**Conclusions:** Pre-sleep brain state determines responsiveness to auditory stimulation, with individuals exhibiting longer sleep onset latency showing greater SWA enhancement. Tailoring pre-sleep interventions based on these predictive features may optimize brain receptivity to auditory stimulation during sleep.

## 1 Introduction

Sleep is a fundamental mental and physical health component, influencing various biological processes[10, 3, 11]. It plays a crucial role in cognitive functions, particularly in memory consolidation and executive functioning[27, 29, 17]. Research shows that inadequate sleep quality leads to cognitive impairments, including deficits in memory, attention, and problem-solving abilities[7, 19], as well as contributing to psychiatric disorders[15, 1]. The interplay between sleep and these conditions is complex; for example, sleep fragmentation significantly impacts cognitive function, particularly in older adults[8, 13].

Sleep is also essential for the brain’s detoxification processes, which help maintain neurological health. During sleep, the brain facilitates the removal of toxic metabolites, including neurotoxic proteins, through the glymphatic system[40]. Dysfunction of sleep could potentially increase the risk for neurodegenerative diseases, as it impairs these detoxification processes[37]. Sleep-related circadian rhythms are crucial for clearing these harmful substances, and disturbances in sleep contribute to their accumulation, leading to cognitive decline and the onset of neurodegenerative conditions[14, 25]. This underscores the importance of maintaining healthy sleep patterns for long-term cognitive health and neurological resilience.

Neural oscillations are crucial for coordinating information processing and facilitating communication among neuronal networks, impacting various cognitive functions including perception, attention, learning, and memory. Notably, slow oscillations (SOs) below 1 Hz are a defining characteristic during slow-wave sleep (SWS)[5]. These oscillations arise from interactions within cortical and thalamic networks, reflecting a synchronized neural activity pattern of alternating states[21]. SOs are instrumental in synaptic downscaling and memory consolidation processes[16], with the synchronization of spindle activities to the up states of SOs being essential for these functions[2] and thought to play a crucial role in memory consolidation and neuroplasticity[28, 12].

Given the functional importance of SOs, research has explored the induction of synchronized cortical SO activity via various external stimulations, including electrical (alternating[18] and direct current[39]), transmagnetic[31], and auditory[38]. However, these methods often impose rhythms irrespective of the brain’s endogenous oscillatory activity, which may contribute to their limited success in enhancing memory retention. Recent advances have seen the development of closedloop systems that employ auditory stimulation in synchrony with the brain’s own rhythms, thereby enhancing SO trains during sleep[23]. The potential for auditory stimulation to modulate sleep dynamics has been further underscored by studies demonstrating its ability to increase the power of delta-frequency Electroencephalogram (EEG) activity[26, 9] and induce K-complexes[24].

The manipulation of sleep dynamics to improve memory performance has been explored through Closed-Loop Auditory Stimulation (CLAS) studies. These studies face limitations in data collection, as they typically require subjects to sleep in sleep centers over multiple nights. To address these constraints, researchers have focused on a stimulation approach that has shown success, specifically the delivery of 50 ms pink noise bursts timed to the upphase of slow oscillations, as demonstrated by Ngo et al.[22]. This method has been shown to prolong a train of previous slow oscillations or increase their amplitude[6], while stimulation during the downtrend can disrupt the oscillatory train[20]. Despite protocol refinements such as precise up-phase timing, variability in individual responses persists, even when stimulus power is adjusted during adaptation through hearing tests. This suggests that the effectiveness of auditory stimulation is closely tied to the individual’s brain state, which can potentially be assessed through pre-sleep measurements, as well as their receptiveness at the time of stimulation.

This study focuses on the relationship between the presleep brain state and responsiveness to auditory stimulation, aiming to predict individual responsiveness using EEG recordings before lights off. The primary objective is to investigate how the pre-sleep brain state influences the effectiveness of CLAS in enhancing slow-wave activity (SWA). We posit that one session of in-lab auditory stimulation targeting slow-wave activity (via tone delivery) enhances memory consolidation and psychomotor functions compared to a sham condition. Furthermore, this study examines the influence of auditory stimulation on sleep-induced changes in both objective and subjective assessments of sleep quality, alertness, and mood. By testing diverse stimulus characteristics and applying stimulation across various sleep stages, this research seeks to clarify the physiological mechanisms through which auditory stimulation modulates sleep architecture and cognitive processes, while providing insights into individual variability in these effects.

## 2 Methods

### 2.1 Participants and Design

Twenty-eight healthy adults (19-40 years, mean *±* SD: 27 *±* 5 years, 16 females, 12 males) with mild sleep restriction participated in this study. All participants were selected following a one-week screening period during which actigraphy and daily sleep logs were used to assess habitual sleep patterns and inclusion criteria. Participants reported regular sleep schedules (bedtime between 9:00 PM and 2:00 AM, wake time before 9:00 AM), sleep latency ≤ 30 minutes, and wake after sleep onset ≤ 30 minutes. They maintained a regular sleep duration of 5-7 hours on work or school nights and extended their sleep by at least one hour on non-work days, either through bedtime extension or daytime napping. They were fluent in English, maintained a stable full-time work or student schedule, and met all inclusion and exclusion criteria (see Appendix A.1). The Northwestern University Institutional Review Board approved this study (Protocol STU00204417), and all participants provided informed consent prior to participation.

Each participant completed three overnight laboratory visits: an adaptation night followed by two experimental nights, separated by 4 *±* 2 days. On one experimental night, auditory stimulation was administered during sleep (Stim), whereas on the other night participants underwent an identical sham condition (Sham) without sound delivery while data and timestamps were recorded. The order of Stim and Sham nights was randomized between participants to minimize order effects.

Each visit consisted of three phases (see Figure 1A): an evening session including cognitive tasks and a presleep baseline recording, an overnight sleep period with polysomnography (PSG) and auditory stimulation or sham. A single input from the midline frontopolar (Fpz) channel was used for online detection of sleep. During the daytime, participants underwent four sessions with repeated cognitive testing and vigilance assessment.

**Figure 1.**
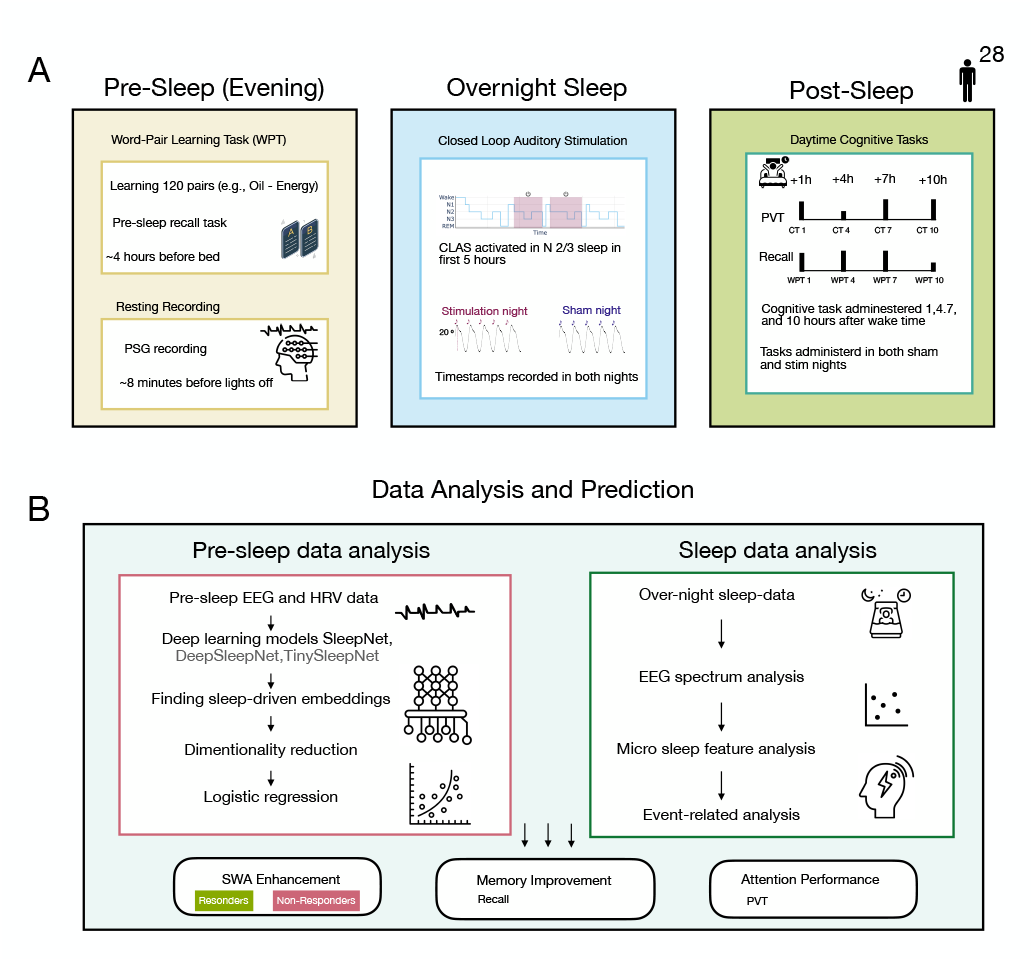
Experimental design and analysis pipeline overview. (**A**) Study protocol spanning three phases. *Pre-Sleep:* Participants (n=28) completed word-pair learning 4h before bed, followed by 8-min resting PSG recording for baseline EEG/HRV. *Overnight Sleep:* Closed-loop auditory stimulation (CLAS) delivered 50ms pink noise bursts timed to slow oscillation upstate during N2/N3 sleep during first 5 hours of sleep. 14 participants had Stim/Sham as their first-non-adaptation-night. Event timestamps recorded for stimulus-locked analyses. *Post-Sleep:* Word recall task and psychomotor vigilance task (PVT) administered at 1, 4, 7, and 11h after wake (CT1, CT4, CT7, CT11) on both nights. (**B**) Analysis pipeline. Pre-sleep EEG/HRV data processed through deep learning models (SleepNet, DeepSleepNet, TinySleepNet) to generate embeddings for logistic regression after dimensionality reduction. Sleep data analyzed for EEG spectral features, slow-wave activity (SWA), spindles, and event-related potentials. Streams converged to predict three outcomes: SWA enhancement (Stim-Sham delta power; responders vs. non-responders by median split), memory improvement, and attention performance.

Participants arrived at the sleep laboratory approximately 5 hours before their habitual bedtime. Following setup of the polysomnographic montage, at least 8 minutes before lights off resting-state recording was acquired to capture baseline EEG and heart rate variability (HRV). The details of protocol explained in Apendix Section A.4. On the two experimental nights (Visits 2 and 3), participants performed a word pair recall task approximately 4 hours before bedtime, as detailed in Appendix section A.3. During this task, participants studied 120 novel word pairs (e.g., oil-energy, tree-river) and attempted cued recall. No items were repeated across sessions, and no feedback was provided during post-sleep testing.

Overnight, participants completed an attended PSG study with continuous monitoring by a trained technologist. The PSG montageincluded EEG, electrooculogram (EOG), submental electromyogram (EMG), and electrocardiogram (ECG) channels, recorded according to American Academy of Sleep Medicine (AASM) standards. Sleep stages were scored manually by a trained scorer following AASM criteria.

Auditory stimulation was delivered using a phase-locked loop (PLL) algorithm that detected and estimated the continous phase of slow oscillations (SOs) [30] (using Fpz channel). Each detected SO triggered a 50 ms burst of pink noise, timed approximately 20^*°*^ before the positive SO peak and repeated five times as shown in Figure. 2A for the first 5 hours of sleep. To assess the immediate effects of stimulation, we used a block-wise alternating protocol with periods of active stimulation (Stim ON) interleaved with control periods (Stim OFF and Sham ON). During Stim ON blocks, auditory bursts were delivered in phase with detected SOs, while during Sham blocks, SOs were detected and triggers were logged, but no sounds were played. Six participants lacking valid trigger timestamps were excluded from time-locked analyses.

**Figure 2.**
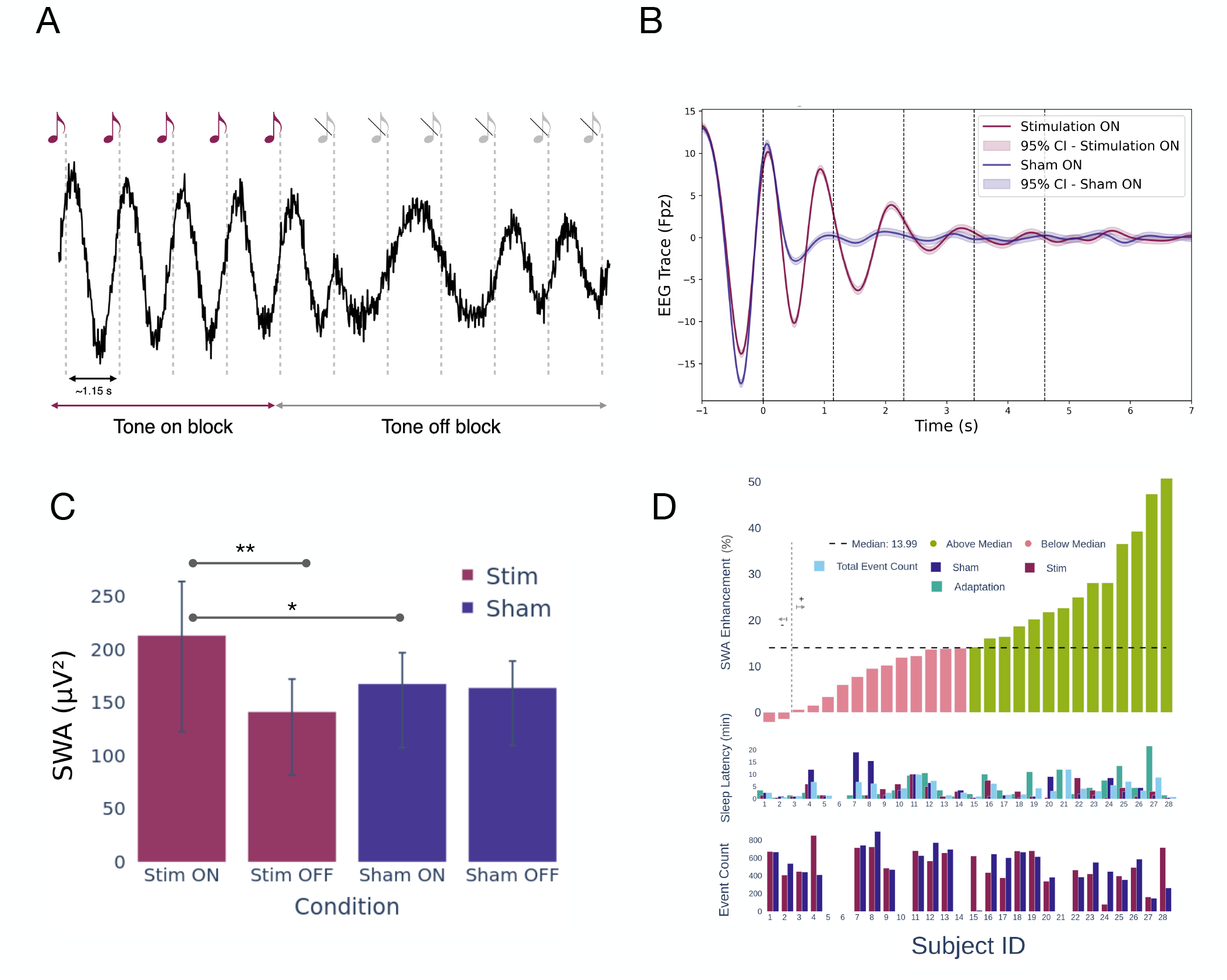
Impact of auditory stimulation on brain activity during sleep. (A) Block-wise Stim–Sham alternation. Each block comprised a fixed number of detected SOs. In Stim blocks, tones were delivered at SO up-state; in Sham blocks, triggers were marked without tones. (B) EEG traces aligned to the first stimulation event (time 0), filtered to 0.25-2Hz and averaged over events for each participnat, then averaged over participants(CI: Confidence Interval). (C) Slow-wave activity (SWA) is higher during Stim ON versus Stim OFF and Sham ON; ^**^p<0.01, ^*^p<0.05. (D) Across-subject representation of SWA enhancement (SWA increase comparing Stim vs Sham). The bottom two ribbons refers to number of times stimulation happened and sleep latency for each night across participants.

Upon awakening, participants repeated cognitive assessments. On each day, participants completed thecognitive tasks at 1, 4, 7, and 10 hours after wake. To assess sustained attention, participants completed the Psychomotor Vigilance Task (PVT). On Visit 1 (adaptation), a 5-minute PVT (administered before recall task) was administered one hour after wake. On Visits 2 and 3, the PVT was administered at 1, 4, 7, and 10 hours after wake.

### 2.2 Stimulation Response Analysis

To assess the impact of stimulation, Wilcoxon rank-sum test were used to compare key features between Stim and Sham nights. Analyses focused on both physiological and behavioral outcomes. For microsleep features, we compared SWA, spindle metrics, and arousal indices across conditions to quantify changes in sleep microstructure. We conducted similar comparison of memory performance and vigilance trajectories between Stim and Sham conditions using recall and PVT.

We split participants by their within-subject change in SWA (Stim minus Sham) using a median split. This is a relative split and it does not imply that every responsive participant had higher SWA on Stim than on Sham. Participants above the median were referred to as responders, and the rest as non-responders. This grouping let us investigate whether EEG and behavioral changes differed between the two.

To visualize and quantify the evoked response to auditory clicks, EEG signals were segmented into 7-second windows spanning −1 s to +6 s relative to each stimulus onset. Grand averages were computed to characterize event-related SO morphology across Stim and Sham conditions.

### 2.3 Pre-Sleep Predictors of Responsiveness

We further investigated whether pre-sleep EEG features could predict responsiveness to CLAS. Using the 8 minutes of pre-sleep recordings (9.58 *±* 0.95 minutes) presleep PSG segment, we extracted spectral features across channels Fpz, F4, F3, C4, C3, P4, P3, O2, O1. Correlation analyses were performed between pre-sleep features and three outcome measures: (1) SWA enhancement, (2) memory recall, and (3) attention performance.

We first tested whether pre-sleep features were associated with these outcomes across participants as shown in Figure 1B. Correlation analyses examined relationships between normalized band power and HRV metrics with the three outcomes defined above: SWA enhancement (Stim minus Sham during ERP windows), recall performance, and sustained attention (median PVT response time). These correlations identified candidate pre-sleep predictors for subsequent machine learning-based analyses.

### 2.4 AI-Driven Prediction

We developed predictive models to predict three outcomes mentioned outcomes (i.e., enhancement of memory, attention, and SWA). Because these outcomes represent continuous variables, we avoided classification at the median and instead focused on discriminating the upper quartile from the lower quartile of responders. This quartile-based approach created balanced classes comparing high performers to low performers while excluding middle-range responses. All models were evaluated using leave-one-subject-out cross-validation to ensure no data leakage between training and testing and to assess generalizability across individuals.

We tested three feature extraction approaches that progressively increased in complexity. The first approach used normalized band power features extracted from presleep EEG as direct inputs. Relative power was calculated for five frequency bands: delta (0.25 to 4 Hz), theta (4 to 8 Hz), alpha (8 to 13 Hz), beta (13 to 30 Hz), and gamma (30 to 40 Hz). Relative power (band power divided by total power) normalized for individual differences in absolute power, yielding five features per recording.

The second and third approaches leveraged transfer learning by extracting embeddings from three pre-trained deep learning architectures: SleepNet [4], DeepSleepNet [32], and TinySleepNet [33]. These models were originally designed to capture temporal dynamics in sleep EEG data and produce 128-dimensional feature vectors that encode complex, non-linear neural patterns.

For the second approach, we extracted embeddings from consecutive temporal windows of the pre-sleep recording, with each of the three models producing 128-dimensional features (384 total features per time step). We then computed mean and standard deviation across time steps, yielding 768 features per recording (384 dimensions × 2 statistics). Principal component analysis (PCA) reduced dimensionality from 768 to 50 components.

The third approach aimed to capture stable, subject-level neural patterns by aggregating embeddings across all recordings for each participant. After extracting embeddings from all recordings and computing temporal statistics (mean and standard deviation) per recording, we averaged these statistics across all recordings for each subject. This created subject-level representations that reduced noise and captured individual differences in neural processing more reliably than single-night recordings. PCA again reduced dimensionality to 50 components.

For each participant, embeddings from both experimental nights (Stim and Sham) were concatenated in a fixed order to ensure consistency across participants. Because recording lengths varied across sessions, sequences were truncated to the minimum common length between conditions before concatenation.

After feature extraction, all three approaches used logistic regression for classifying patterns. For bandpower features, models were applied directly without dimensionality reduction. For embedding approaches, PCA preprocessing preceded model training. All models predicted binary responsiveness labels (upper quartile vs lower quartile) using leave-one-subject-out cross-validation. Computational details and software versions are provided in Appendix A.5.

## 3 Results

### 3.1 Auditory Stimulation Enhances Slow-Wave Activity

Closed-loop auditory stimulation increased slow-wave activity during NREM sleep. Event-related potential analysis showed enhancement of slow oscillations time-locked to auditory clicks (Figure 2B). This enhancement was most pronounced in the delta frequency range (0.5-4 Hz), as shown in the time-frequency analysis in Appendix Section A.2.

Delta power during tone-on blocks averaged 213.4 *±* 126.5 *µ*V^2^, significantly higher than during tone-off blocks (141.5 *±* 90.5 *µ*V^2^). In contrast, sham nights showed no difference between tone-on (167.9 *±* 102.6 *µ*V^2^) and tone-off periods (164.2 *±* 96.7 *µ*V^2^) (Figure 2C). This pattern indicates that stimulation concentrated slow-wave activity into targeted windows, with a corresponding reduction during off periods. To investigate whether stimulation efficacy varied across the night, we compared SWA between

Stim and Sham conditions for each sleep cycle. Because stimulation was delivered only during the first 5 hours of sleep, we expected SWA differences to be concentrated in early cycles, with convergence between conditions in later cycles after stimulation stoped. As shown in Figure 3, SWA during Stim nights was significantly elevated compared to Sham nights during the first sleep cycle (201.3 *±* 108.5 *µ*V^2^ vs. 183.6 *±* 112.8 *µ*V^2^; *p* < 0.05) and the second sleep cycle (198.7 *±* 98.4 *µ*V^2^ vs. 142.1 *±* 98.7 *µ*V^2^; *p* < 0.05). The most robust difference emerged during the third sleep cycle (163.2 *±* 48.6 *µ*V^2^ vs. 76.8 *±* 56.3 *µ*V^2^; *p* < 0.01), suggesting that the cumulative effects of stimulation were particularly pronounced during this period. However, by the fourth and fifth sleep cycles, when slow-wave sleep naturally declines and no stimulation had been delivered, the differences between Stim and Sham conditions were no longer statistically significant.

**Figure 3.**
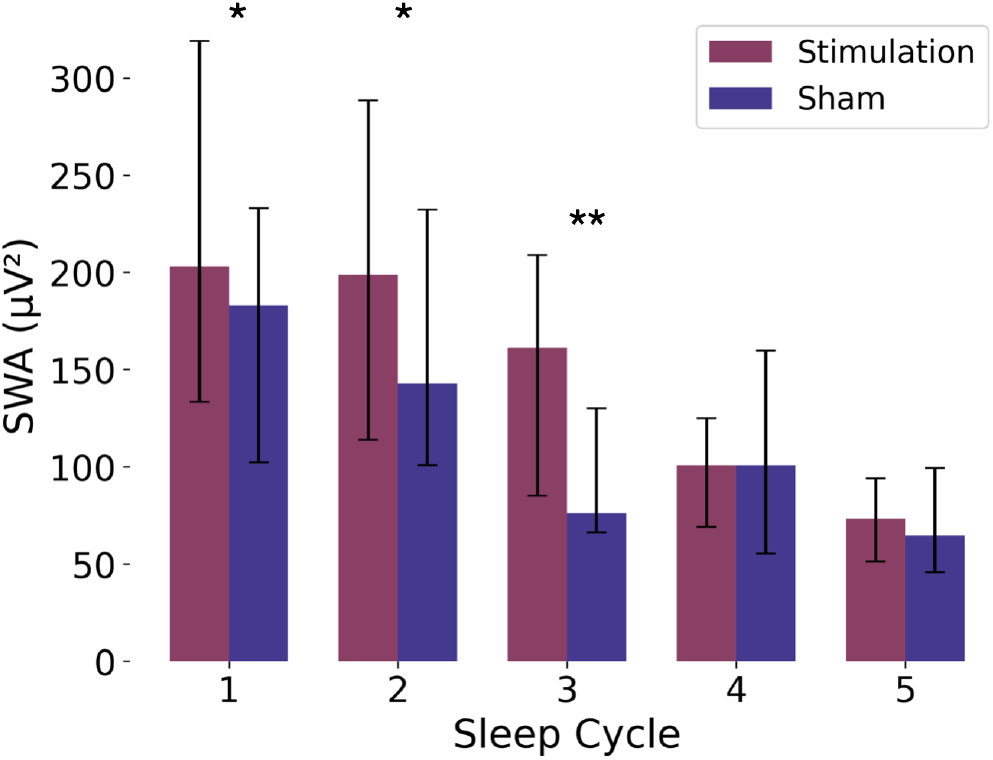
Slow-wave activity enhancement across sleep cycles. Comparison of SWA between stimulation and sham nights for each sleep cycle. Error bars indicate standard deviation. ^**^p < 0.01, ^*^p < 0.05.

### 3.2 Inter-Pulse Timing Within Stimulation Bursts

To assess the temporal precision of the stimulation system, we quantified the inter-stimulus intervals within each five-pulse click burst (Δ1–Δ4). These Δ values represent the latency between successive clicks inside a burst (e.g., Δ_1_ is the time between click 1 and click 2). Interpulse intervals were averaged per participant, and group-level metrics were computed. As shown in Table 1, interpulse timing was highly consistent across pulse positions, with minimal variability across subjects. Across the cohort, the mean inter-click interval was 1.166 *±* 0.028 s (mean *±* SD).

**Table 1:** Inter-pulse intervals within stimulation bursts (mean and standard deviation in seconds).

| Pulse interval | $\Delta 1$ | $\Delta 2$ | $\Delta 3$ | $\Delta 4$ |
| --- | --- | --- | --- | --- |
| Mean (s) | 1.159 | 1.167 | 1.170 | 1.167 |
| SD (s) | 0.029 | 0.030 | 0.031 | 0.025 |

### 3.3 Interindividual Variability In Responsiveness

Participants’ responses showed substantial heterogeneity in SWA enhancement (Figure 2D). The top panel displays the percentage change in SWA for each participant, with subjects ranked by their degree of enhancement. Twentysix out of twenty-eight participants showed an increase in SWA, with the dashed line indicating the median level of enhancement. The middle and bottom panels illustrate the raw event counts for total, sham, and stim conditions across subjects, demonstrating that the observed effects were not driven by differences in the number of stimulation events. For six participants, although the closed-loop system operated correctly, stimulus timestamps were not recorded in the EEG trigger channel, as shown in the bottom panel of Figure 2D. This variability in responsiveness motivated subsequent analyses to identify predictors of individual differences in stimulation efficacy.

Sleep architecture remained unchanged by stimulation. Sleep efficiency, total sleep time, stage distributions, arousal indices, and spindle characteristics were similar between stimulation and sham nights (Table 2, all p > 0.05). Arousals were quantified using a stage-transition approach that identified shifts from stable NREM sleep to wakefulness or lighter stages and from REM sleep to wake/N1, yielding overall and stage-specific arousal indices. Spindle events were automatically detected using the open-source YASA library [35] applied to the Fpz EEG channel. More details on methodology used are provided in Section A.9.

**Table 2:**
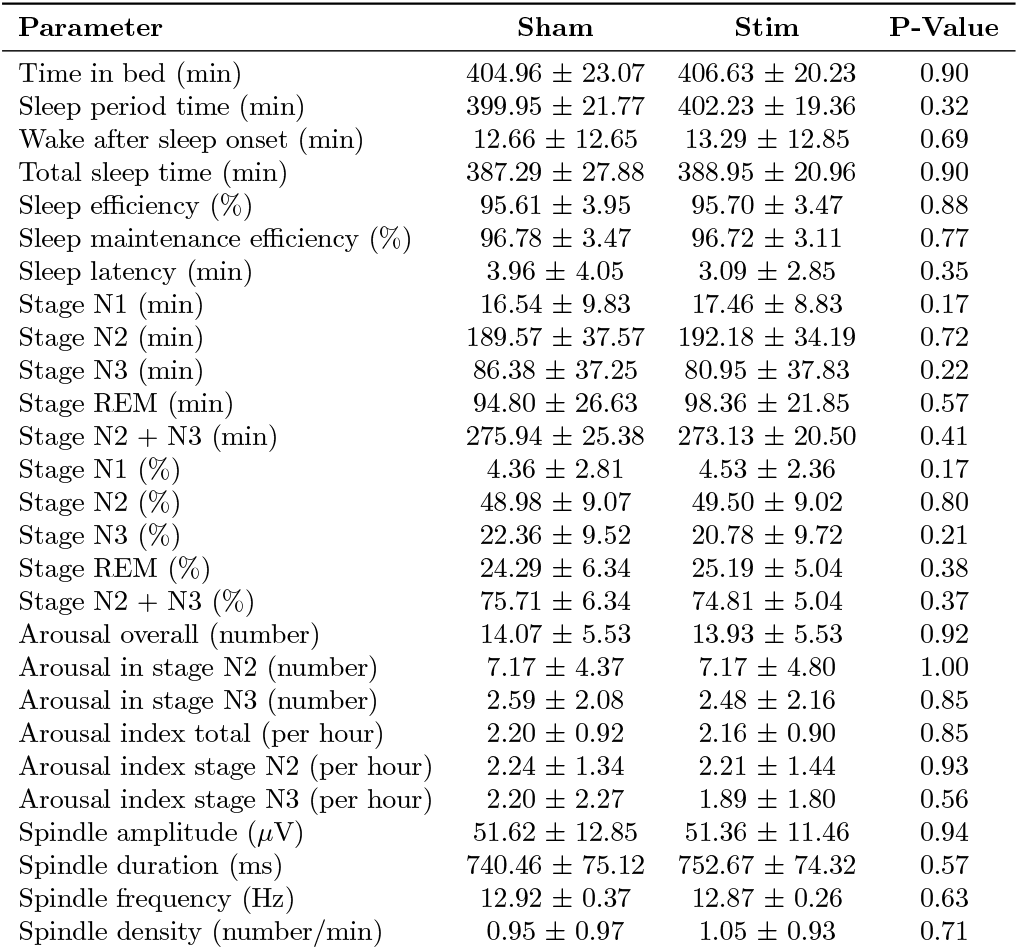
Sleep macrostructure and spindle characteristics for stim and sham nights.

| Parameter | Sham | Stim | P-Value |
| --- | --- | --- | --- |
| Time in bed (min) | 404.96 ± 23.07 | 406.63 ± 20.23 | 0.90 |
| Sleep period time (min) | 399.95 ± 21.77 | 402.23 ± 19.36 | 0.32 |
| Wake after sleep onset (min) | 12.66 ± 12.65 | 13.29 ± 12.85 | 0.69 |
| Total sleep time (min) | 387.29 ± 27.88 | 388.95 ± 20.96 | 0.90 |
| Sleep efficiency (%) | 95.61 ± 3.95 | 95.70 ± 3.47 | 0.88 |
| Sleep maintenance efficiency (%) | 96.78 ± 3.47 | 96.72 ± 3.11 | 0.77 |
| Sleep latency (min) | 3.96 ± 4.05 | 3.09 ± 2.85 | 0.35 |
| Stage N1 (min) | 16.54 ± 9.83 | 17.46 ± 8.83 | 0.17 |
| Stage N2 (min) | 189.57 ± 37.57 | 192.18 ± 34.19 | 0.72 |
| Stage N3 (min) | 86.38 ± 37.25 | 80.95 ± 37.83 | 0.22 |
| Stage REM (min) | 94.80 ± 26.63 | 98.36 ± 21.85 | 0.57 |
| Stage N2 + N3 (min) | 275.94 ± 25.38 | 273.13 ± 20.50 | 0.41 |
| Stage N1 (%) | 4.36 ± 2.81 | 4.53 ± 2.36 | 0.17 |
| Stage N2 (%) | 48.98 ± 9.07 | 49.50 ± 9.02 | 0.80 |
| Stage N3 (%) | 22.36 ± 9.52 | 20.78 ± 9.72 | 0.21 |
| Stage REM (%) | 24.29 ± 6.34 | 25.19 ± 5.04 | 0.38 |
| Stage N2 + N3 (%) | 75.71 ± 6.34 | 74.81 ± 5.04 | 0.37 |
| Arousal overall (number) | 14.07 ± 5.53 | 13.93 ± 5.53 | 0.92 |
| Arousal in stage N2 (number) | 7.17 ± 4.37 | 7.17 ± 4.80 | 1.00 |
| Arousal in stage N3 (number) | 2.59 ± 2.08 | 2.48 ± 2.16 | 0.85 |
| Arousal index total (per hour) | 2.20 ± 0.92 | 2.16 ± 0.90 | 0.85 |
| Arousal index stage N2 (per hour) | 2.24 ± 1.34 | 2.21 ± 1.44 | 0.93 |
| Arousal index stage N3 (per hour) | 2.20 ± 2.27 | 1.89 ± 1.80 | 0.56 |
| Spindle amplitude (μV) | 51.62 ± 12.85 | 51.36 ± 11.46 | 0.94 |
| Spindle duration (ms) | 740.46 ± 75.12 | 752.67 ± 74.32 | 0.57 |
| Spindle frequency (Hz) | 12.92 ± 0.37 | 12.87 ± 0.26 | 0.63 |
| Spindle density (number/min) | 0.95 ± 0.97 | 1.05 ± 0.93 | 0.71 |

### 3.4 Cognitive Performance

#### 3.4.1 Memory Performance

When examining memory improvement relative to baseline (Figure 4A), performance declined progressively throughout the day in both conditions. A linear trend analysis across all time points showed a significant negative correlation with time for stimulation (*r* = −0.39, *p* < 0.001), sham (*r* = −0.38, *p* < 0.001), and when averaged across conditions (*r* = −0.46, *p* < 0.001). The magnitude and rate of this time-dependent decline were comparable between conditions, indicating that stimulation did not alter the temporal trajectory of recall performance.

**Figure 4.**
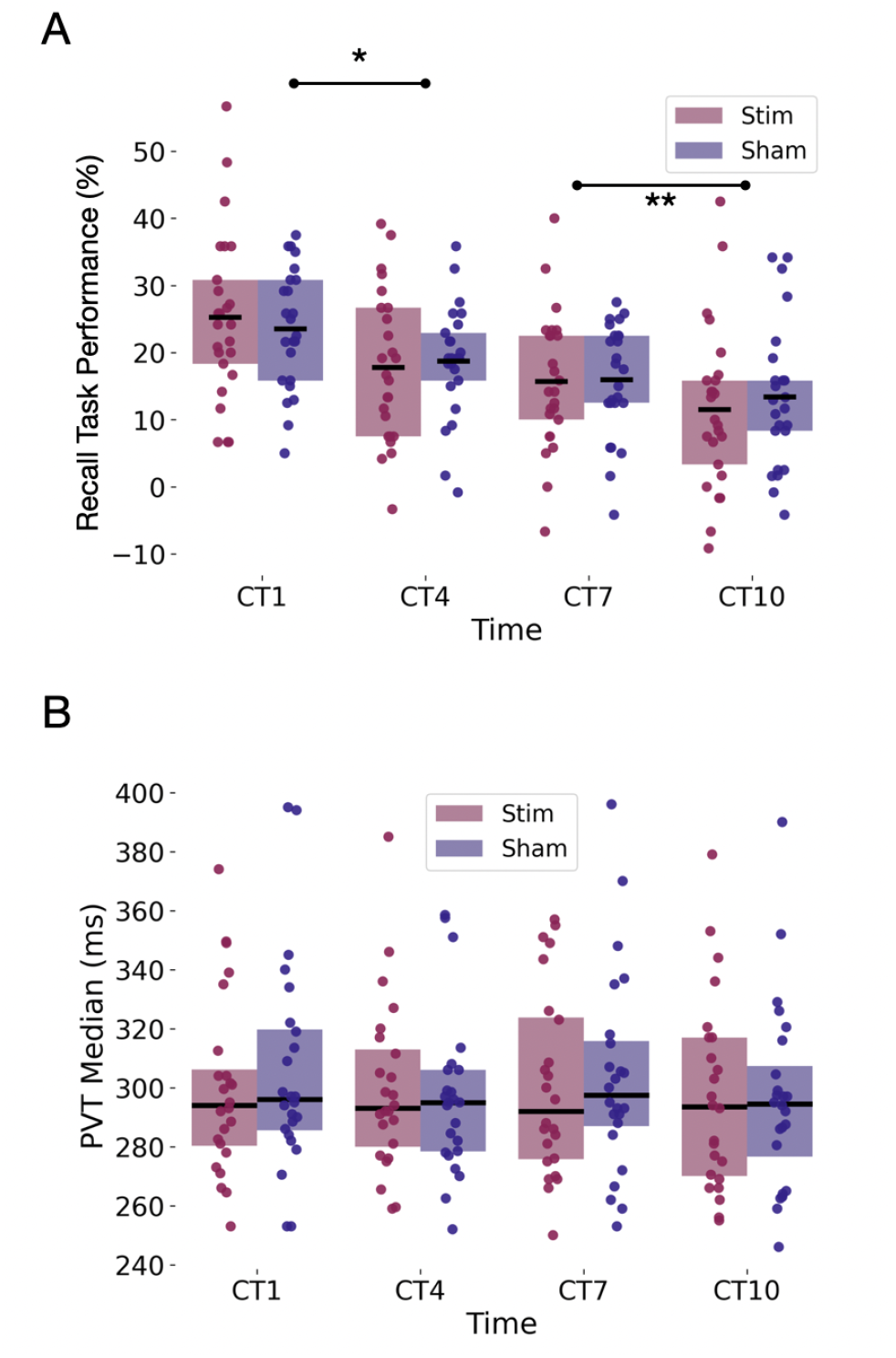
Cognitive tasks (CT) across circadian time points. (A) Shows the memory recall performance compared to the baseline (pre-sleep) in 1, 4, 7, and 10 hours after wake time. Box plots display the median (center line), interquartile range (box), with individual data points overlaid. (B) Psychomotor vigilance Task (PVT) median response times.

#### 3.4.2 Sustained Attention

Psychomotor vigilance task performance remained stable across the day with no effect of stimulation (Figure 4B). Median response times were comparable between conditions at all timepoints. Response time variability was also similar between conditions across all measurement points. The combined analysis showed no significant changes in vigilance across the day (all comparisons p > 0.05), indicating that neither condition produced deterioration in sustained attention performance.

This suggests while auditory stimulation enhanced SWA during sleep, this enhancement did not translate to measurable improvements in next-day recall performance or sustained attention under the current protocol (i.e,. CLAS with pink noise) while we aggregate the results for the whole population.

Given the lack of overall effects, we studied whether cognitive outcomes differed between individuals who showed physiological responsiveness to stimulation (i.e., SWA enhancement) and those who did not.

#### 3.4.3 Performance by Physiological Response

Participants were classified as responders or non-responders based on SWA enhancement. At 4 hours post-wake, responders showed better memory retention than non-responders (median improvement: 3.3% vs −4.5%, p = 0.07). At 7 hours, the difference persisted (4.2% vs −3.3%, p = 0.08). No differences appeared at 1 hour (p = 0.26) or 10 hours (p = 0.68) (Figure 5A). Also, sustained attention did not differ between responders and non-responders at any timepoint (all p > 0.27) (Figure 5B).

**Figure 5.**
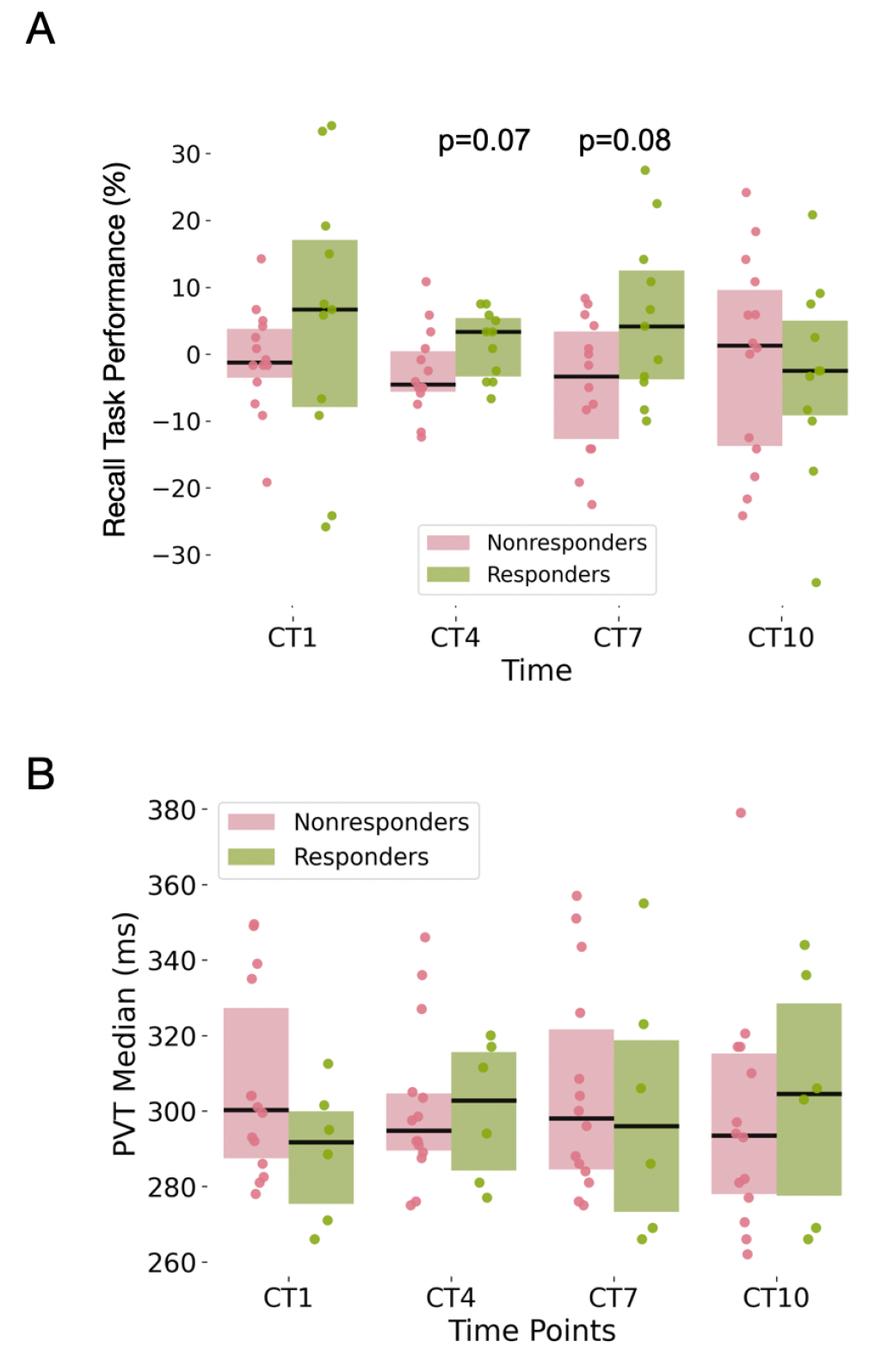
Cognitive outcomes in responders versus non-responders. (A) Word-pair test score improvement (Stim minus Sham, relative to baseline) across circadian timepoints. Responders (green) showed trends toward better memory retention at CT4 (p = 0.07) and CT7 (p = 0.04) compared to non-responders (pink). (B) Psychomotor vigilance task median response times across circadian timepoints. No significant differences were observed between responders and non-responders at any timepoint.

We conducted a repeated-measures ANOVA to examine how recall performance varied across time and condition. We used the change from baseline (Post −Pre) at each of the four time points (1, 4, 7, 10 hours). Separately for non-responders and responders, we ran a 4 *×* 2 repeated-measures ANOVA with within-subject factors Time (four post-intervention time points) and Condition (Sham vs Stim). Non-responders showed a large main effect of Time, *F* (3, 39) = 11.36, *p* = 1.71 *×* 10^−5^, 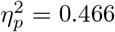, no main effect of Condition, *F* (1, 13) = 1.60, *p* = 0.228, 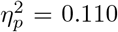, and no Time *×* Condition interaction, *F* (3, 39) = 0.57, *p* = 0.636, 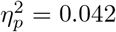. Responders similarly showed a large Time effect, *F* (3, 30) = 8.24, *p* = 3.78 × 10^−4^, 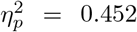, no Condition effect, *F* (1, 10) = 0.70, *p* = 0.422, 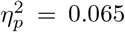, and no interaction, *F* (3, 30) = 1.08, *p* = 0.372, 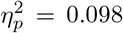. These results indicate that recall performance changed significantly over the 10-hour period in both groups, but there was no evidence that Stim versus Sham conditions differed overall or showed different temporal patterns.

### 3.5 Pre-Sleep Predictors of Responsiveness

#### 3.5.1 EEG Spectral Power and SWA Enhancement

As it is shown in Figure A5, pre-sleep alpha power correlated with SWA enhancement across adaptation (*r* = 0.413, *p* = 0.04), stimulation (*r* = 0.435, *p* = 0.04), sham (*r* = 0.542, *p* = 0.01), and combined sessions (*r* = 0.436, *p* < 0.01). Theta power showed positive correlations in combined analysis (*r* = 0.329, *p* < 0.01). Delta power showed negative correlations in adaptation (*r* = −0.360, *p* = 0.08) and combined data (*r* = −0.299, *p* = 0.01).

#### 3.5.2 EEG Power and Cognitive Outcomes

Figure A6 shows that pre-sleep alpha power correlated negatively with memory improvement at 7 and 10 hours post-sleep (combined analysis: *p* = 0.07, *p* = 0.03). Gamma power correlated positively with memory at multiple timepoints (*p* = 0.04 to *p* = 0.01).

For sustained attention, delta and theta bands correlated with PVT performance at 1 hour (delta: *p* < 0.01; theta: *p* < 0.01) and 7 hours (delta: *p* = 0.03; theta: *p* = 0.03) in combined analysis (see Figure 6).

**Figure 6.**
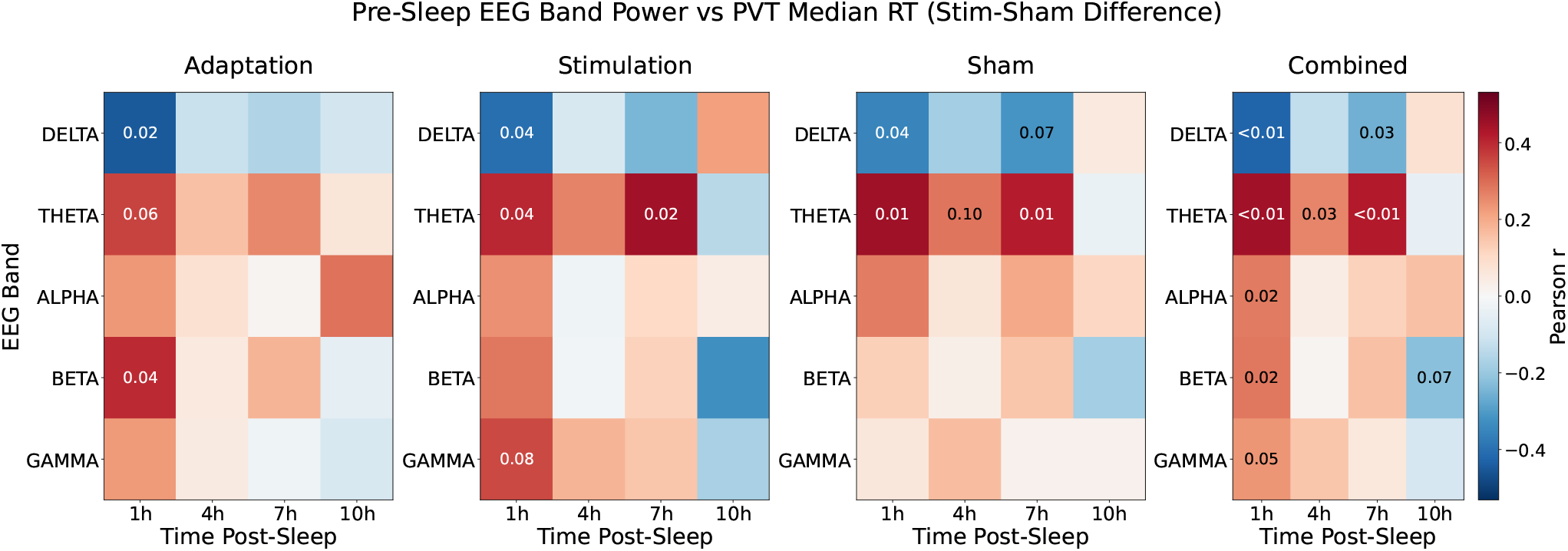
Correlation between pre-sleep EEG band power and psychomotor vigilance test performance differential (Stim-Sham) across post-sleep time points. Heatmaps showing Pearson correlation coefficients between normalized pre-sleep EEG spectral power in different frequency bandsand the difference in PVT median reaction time between stimulation and sham conditions at 1, 4, 7, and 10 hours post-sleep for adaptation,

#### 3.5.3 Sleep Onset Latency

Figure 7 represents sleep onset latency correlation with SWA enhancement across adaptation (*r* = 0.424, *p* = 0.03), stimulation (*r* = 0.479, *p* = 0.01), sham (*r* = 0.465, *p* = 0.02), and combined data (*r* = 0.429, *p* < 0.01).

**Figure 7.**
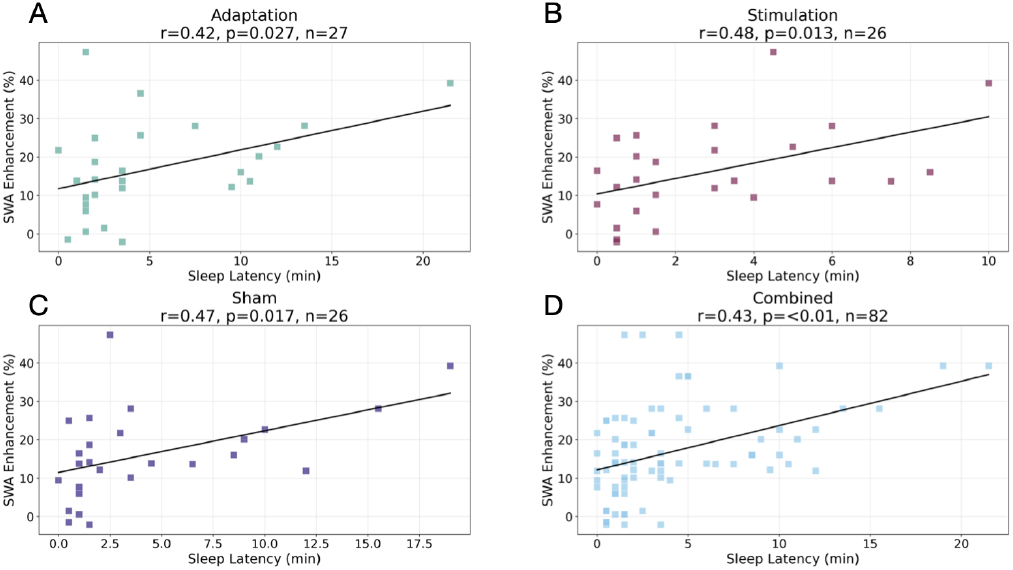
Sleep latency predicts SWA enhancement. Correlation between sleep onset latency and SWA enhancement across session types. Each point represents one participant night.

#### 3.5.4. Stimulation Event Count

Number of stimulation events correlated negatively with SWA enhancement in stimulation (*r* = −0.37, *p* = 0.086), sham (*r* = −0.40, *p* = 0.056), and combined data (*r* = −0.47, *p* = 0.023) (see Figure 8).

**Figure 8.**
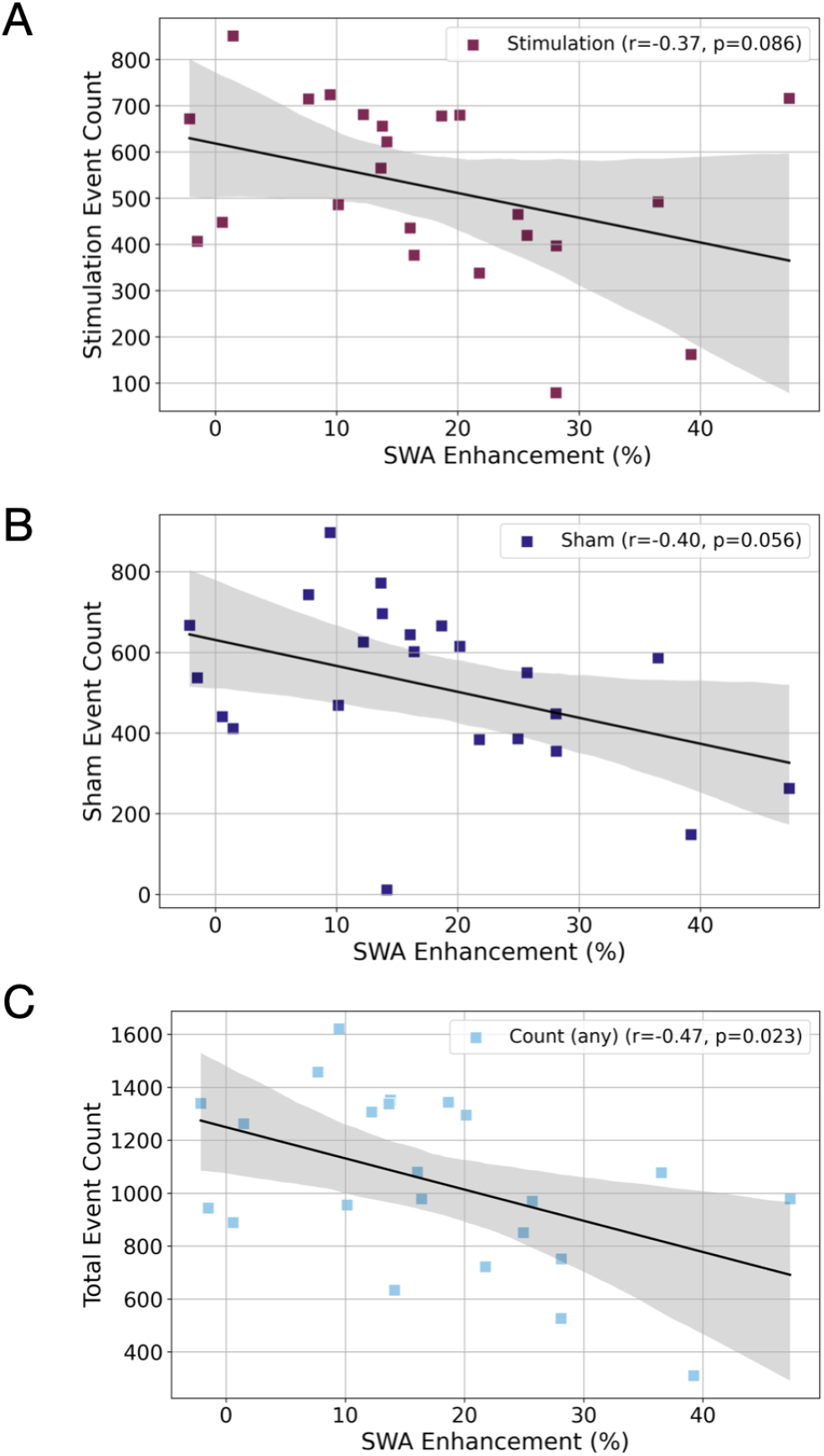
Fewer stimulation opportunities predict greater enhancement. Correlation between stimulation event count and SWA enhancement for (A) stimulation nights, (B) sham nights, and (C) combined data. Each point represents one participant night.

### 3.6 Prediction of Responsiveness

#### 3.6.1. Early Response Prediction

Ridge regression models trained on spectral features from the first five ERP windows (7 seconds each) predicted night-long responsiveness (MSE p = 0.0858, MAE p = 0.0449). Predictive features concentrated in slow-wave (1-5 Hz) and theta (4-8 Hz) bands during 0-6 seconds post-stimulus (see Figure 9).

**Figure 9.**
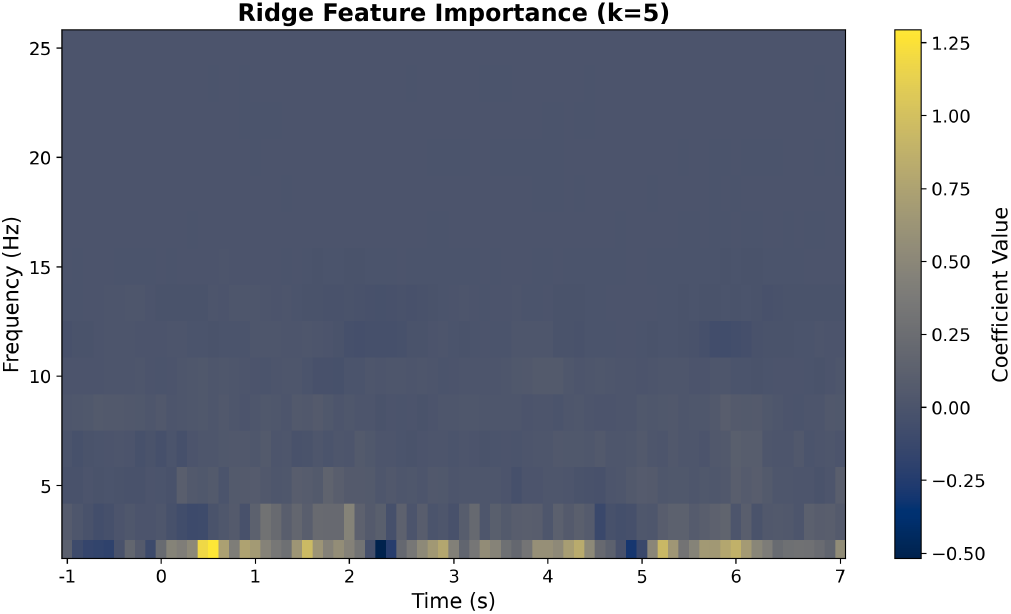
Early neural responses predict night-long enhancement. Time-frequency representation of ridge regression coefficients for predicting SWA enhancement from the first five ERP windows. Warmer colors indicate positive associations with enhancement, cooler colors indicate negative associations.

#### 3.6.2 Pre-Sleep Feature Classification

Classification models achieved varying accuracy across feature types (Table 3). For SWA enhancement: band-power (72% accuracy, AUC 0.66), night-level embeddings (72% accuracy, AUC 0.86), subject-level embeddings (80% accuracy, AUC 0.93). For memory enhancement: subject-level embeddings achieved 90% accuracy (AUC 0.92). For attention: bandpower achieved 64% accuracy (AUC 0.68).

**Table 3:** Prediction performance for SWA enhancement, Memory enhancement (ΔRecall adj.), and Attention improvement (ΔPVT) across feature sets.

| Feature Set | Target | Acc | Prec | Recall | Spec | F1 | AUC |
| --- | --- | --- | --- | --- | --- | --- | --- |
| Bandpower | SWA Enhancement | 0.72 | 0.76 | 0.65 | 0.80 | 0.70 | 0.66 |
| Night-level Embedding | SWA Enhancement | 0.72 | 0.74 | 0.70 | 0.75 | 0.72 | 0.86 |
| Subject-level Embedding | SWA Enhancement | 0.80 | 0.83 | 0.71 | 0.88 | 0.77 | 0.93 |
| Bandpower | Memory Enhancement (ΔRecall adj.) | 0.65 | 0.65 | 0.79 | 0.50 | 0.71 | 0.73 |
| Night-level Embedding | Memory Enhancement (ΔRecall adj.) | 0.77 | 0.79 | 0.79 | 0.75 | 0.79 | 0.74 |
| Subject-level Embedding | Memory Enhancement (ΔRecall adj.) | 0.90 | 1.00 | 0.80 | 1.00 | 0.89 | 0.92 |
| Bandpower | Attention Improvement (ΔPVT) | 0.64 | 0.65 | 0.61 | 0.67 | 0.63 | 0.68 |
| Night-level Embedding | Attention Improvement (ΔPVT) | 0.58 | 0.67 | 0.33 | 0.83 | 0.44 | 0.61 |
| Subject-level Embedding | Attention Improvement (ΔPVT) | 0.54 | 0.57 | 0.57 | 0.50 | 0.57 | 0.50 |

## 4 Discussion

This study examined individual differences in responsiveness to closed-loop auditory stimulation during sleep and investigated whether pre-sleep physiological features could predict these differences. Our findings demonstrate three main results: (1) auditory stimulation successfully enhanced SWA during NREM sleep, with effects concentrated in the first three sleep cycles when stimulation was delivered; (2) substantial inter-individual variability existed in responsiveness, with some participants showing marked SWA enhancement while others showed minimal or no response; and (3) pre-sleep brain state, particularly spectral power and heart rate variability, predicted responsiveness to stimulation. Individuals who responded to stimulation also showed modest benefits in memory consolidation during the mid-day period following sleep.

### 4.1 Enhanced Response in High-Need Individuals

The relationship between sleep onset latency and SWA enhancement reveals who benefits most from stimulation (Figure 7). Participants who took longer to fall asleep showed greater SWA enhancement. This pattern appeared across adaptation, stimulation, and sham nights, indicating it reflects a fundamental property of sleep regulation rather than a stimulation-specific effect. Delayed sleep onset may indicate higher pre-sleep arousal that, once sleep is achieved, creates conditions favorable for being more receptive to CLAS with pink noise.

The negative correlation between stimulation event count and SWA enhancement provides complementary insight (Figure 8). This relationship appeared in both stimulation and sham nights, ruling out the interpretation that more stimulation causes diminishing returns. Instead, because the closed-loop algorithm delivers stimulation only when detecting suitable slow oscillations, fewer events indicate fewer opportunities for stimulation. These individuals with weaker endogenous slow-wave generation showed greater enhancement when stimulation was delivered. Together with the sleep latency findings, this suggests that individuals with higher arousal or weaker baseline slow-wave activity respond best to auditory stimulation.

### 4.2 Multi-Night Recordings Improve Prediction Accuracy

Machine learning models classified participants as responders or non-responders using three feature sets: spectral bandpower, night-level embeddings, and subject-level embeddings (Table 3). Subject-level embeddings, which aggregate patterns across multiple nights, consistently outperformed single-night features for predicting SWA and memory enhancement. For attention outcomes, prediction accuracy remained low across all feature sets.

The superiority of subject-level embeddings indicates that stable individual differences in neural processing determine responsiveness more than night-to-night variations. By averaging across adaptation, stimulation, and sham nights, this approach captures consistent individual patterns while reducing noise. For clinical applications, this finding suggests that multiple baseline recordings provide more reliable prediction than single-night assessments. The subject-level approach offers a method to identify individuals most likely to benefit from sleep enhancement interventions before beginning treatment.

### 4.3 Toward Optimized and Personalized Auditory Stimulation

The observation that pre-sleep brain state drives responsiveness suggests a novel two-phase approach to auditory stimulation. Rather than only delivering stimulation during sleep, interventions could begin with a pre-sleep phase designed to bring the brain into a receptive state. This preparatory phase might use acoustic stimulation, sensory entrainment, or other non-invasive techniques to modulate arousal levels, or prime slow-wave generating circuits. Once the brain reaches an optimal state, as assessed by real-time monitoring of spectral features or other biomarkers, the system could transition to phaselocked stimulation during sleep.

This framework represents a departure from current approaches that treat all individuals and all nights identically. By acknowledging that responsiveness varies systematically with pre-sleep state, we can begin to design adaptive interventions that optimize both the preparatory conditions and the stimulation protocol itself. Machine learning models trained on pre-sleep features provide the foundation for such systems, enabling real-time prediction of responsiveness and dynamic adjustment of intervention parameters.

The results of this study represent an important first step in this direction by establishing that pre-sleep features contain predictive information about responsiveness and that early neural responses can validate these predictions within minutes of sleep onset. Future work should investigate which specific pre-sleep interventions most effectively enhance receptivity to closed-loop stimulation and whether optimizing both phases produces greater and more consistent benefits than stimulation alone.

### 4.4 Cognitive Outcomes and the Role of Individual Differences

The relationship between SWA enhancement and cognitive outcomes was modest and selective. At the group level, stimulation did not improve recall performance or sustained attention compared to sham. However, when participants were separated by responsiveness, individuals who showed physiological enhancement of SWA also demonstrated trends toward better memory retention during the mid-day period (4-7 hours after wake). This suggests that cognitive benefits of slow-wave enhancement may be restricted to individuals who achieve sufficient physiological response.

### 4.5 Conclusion

This study demonstrates that responsiveness to closedloop auditory stimulation varies systematically with presleep brain state, enabling prediction of who will benefit most from enhancement. Pre-sleep alpha and theta band power predicted overnight SWA enhancement, while different frequency bands (alpha, gamma, delta, theta) predicted distinct cognitive outcomes including memory consolidation and sustained attention. Also individuals with longer sleep onset latency showed greater SWA enhancement (r = 0.429, p < 0.01), indicating that those who need enhancement most benefit most from stimulation. Machine learning models achieved 64 to 80% accuracy predicting responsiveness from pre-sleep spectral features, with transfer learning embeddings substantially improving discrimination. Temporal embedding features achieved the highest performance (AUC = 0.93 for SWA prediction, AUC = 0.92 for memory enhancement), demonstrating that dynamic patterns in pre-sleep activity provide superior predictive information. This prediction capacity enables personalized stimulation protocols tailored to individual pre-sleep states, adaptive closed-loop systems that assess responsiveness in real time and adjust parameters accordingly, and targeted interventions for individuals most likely to benefit. The findings support a two-phase framework that first optimizes pre-sleep conditions to enhance brain receptivity, then delivers closed-loop stimulation with parameters adjusted based on early neural responses. By identifying pre-sleep biomarkers of responsiveness and demonstrating their link to both physiological and cognitive outcomes, this work establishes the foundation for precision approaches to sleep enhancement that move beyond one size fits all protocols toward interventions optimized for individual physiological readiness.

## 6 Data Availability

All data produced in the present study are available upon reasonable request to the authors.

## 7 Acknowledgments

YK, AL, and GC acknowledge support from the Alzheimer’s Association, The Michael J. Fox Foundation for Parkinson’s Research, and CurePSP Sleep Contributions to Neurodegeneration Grant Program (SCN25-1470707).

## A Appendix

### A.1 Exclusion Criteria

Participants were excluded if they had major medical or psychiatric conditions; current or recent use of sleep medications, stimulants, or psychoactive drugs; body mass index > 30 kg/m^2^; diagnosed sleep disorders (obstructive sleep apnea, restless legs syndrome, insomnia, parasomnias, circadian rhythm disorders); high risk of sleep apnea based on STOP-BANG or Berlin questionnaires; moderate hearing loss; seizure history; recent drug or alcohol abuse; or excessive daily caffeine use (> 500 mg). Participants with alpha–delta sleep on the first laboratory night or drastic changes in sleep duration prior to any visit were also excluded.

### A.2 Time-Frequency Analysis of Stimulation Effects

To further characterize the frequency-specific effects of auditory stimulation, we examined time-frequency dynamics using spectrogram analysis (Figure A1). During Stimconditions (Figure A1A), tone delivery marked by vertical dashed lines was followed by pronounced enhancement of low-frequency power, particularly in frequencies below 5 Hz. This enhancement was most prominent in the delta band (0.5–4 Hz) and appeared within the first second following tone onset, persisting for several seconds. In contrast, Sham conditions (Figure A1B) showed no comparable modulation of spectral power at the same timing markers, despite identical slow oscillation detection and trigger placement. The difference spectrogram (Stim-Sham; Figure A1C) directly quantified this differential response, revealing that auditory stimulation specifically enhanced delta-band power shortly after tone onset, with peak differences reaching up to 10 dB. This time-frequency analysis confirms that the observed neural response was both frequency-specific and temporally locked to the auditory stimulus.

### A.3 Recall task through word pair test

#### A.3.1 Night Study

Participants first completed a study phase to encode word associations. On each trial, a pair of words appeared vertically on the screen (cue on top, associate below) for 4 s. A total of 120 unique pairs were presented, each exactly once. Participants were instructed to pay close attention and try to memorize each pair for a later test.

#### A.3.2 Night Learning (Practice)

Immediately after the study phase, participants practiced cued recall with feedback. On each trial, only the cue word (the top word from study) was shown above a blank line. Participants: 1. Pressed the space bar as soon as they recalled the associate. 2. Typed the associate and pressed Enter to submit. 3. Viewed the correct pair, which was then displayed as feedback. Participants were encouraged to enter their best guess when uncertain; if they had no idea, they could submit a blank response by pressing Enter. Each pair was practiced twice, with a scheduled break halfway through the practice set.

#### A.3.3 Day Testing

Delayed cued recall was administered during daytime sessions labeled. Trial mechanics matched the practice phase without feedback: 1. The cue word appeared above a blank line. 2.Participants pressed the space bar upon recall, typed the associate, and pressed Enter. Only one response was permitted per pair. Participants were instructed to provide their best guess when unsure and to focus carefully on each trial.

### A.4 Presleep HRV Baseline

Recordings were acquired with BrainVision Recorder. Before acquisition, electrode impedances were checked and kept below 5 kΩ when possible, with values below 10 kΩ accepted. The montage was verified and standard system calibrations were performed.

Participants lay in a supine position with hands by their sides and eyes open. They were instructed to remain still and to keep their eyes open for the entire recording. A continuous 8 min baseline was collected for heart rate variability analysis.

A short biocalibration sequence was recorded for artifact labeling and quality control. The sequence included eyes open, eyes closed, upward gaze, downward gaze, leftward gaze, rightward gaze, five blinks, and a brief jaw clench.

### A.5 Computational Resources and Software

All analyses were performed in Python 3.9 using PyTorch 2.0 (CUDA 11.8) for deep learning, and scikitlearn 1.3 for machine learning and feature processing. Data manipulation and statistical analyses utilized NumPy 1.24, Pandas 2.0, and SciPy 1.11. All experiments were executed on a workstation equipped with an NVIDIA GPU (16 GB VRAM). To ensure reproducibility, random seeds were fixed for cross-validation splitting, model initialization, and data shuffling operations.

### A.6 Investigating the Correlation of Heat Rate Variability and Cognitive Outcomes

We examined whether pre-sleep cardiac autonomic activity, indexed by heart rate variability (HRV) metrics, could predict cognitive outcomes. Figure A4 displays correlations between various HRV metrics (pNNx measures, representing the percentage of successive normal-to-normal intervals differing by more than x milliseconds) and changes in word-pair test and PVT performance. Despite examining multiple HRV indices and timepoints, no consistent or statistically significant relationships emerged between pre-sleep HRV and subsequent cognitive performance changes. Only sporadic weak correlations with *p* ≤ 0.1 were observed, with no coherent pattern across metrics or timepoints. This suggests that, unlike pre-sleep EEG features, cardiac autonomic activity measured before sleep does not reliably predict individual differences in cognitive responses to auditory stimulation.

### A.7 Participant Demographics

Twenty-eight participants (16 females, 12 males) completed the study, with a mean age of 27.1 *±* 4.8 years (range: 19 to 37 years). The sample was ethnically and racially diverse, with approximately one-third identifying as Hispanic or Latino and representation across multiple racial categories. Mean body mass index was 25.1 *±* 2.9 kg/m^2^, within the normal to slightly overweight range. Most participants had completed four or more years of college (85.7%), were single (82.1%), and were employed (89.3%). Work and school commitments varied, with approximately 43% engaged full-time in work, 43% engaged full-time in school, and the remainder engaged part-time or not at all. This sample represents a typical young adult population experiencing mild sleep restriction due to work or academic demands. Complete demographic characteristics are presented in Table A1.

### A.8 Heart Rate Variability Measures

The HRV metrics were extracted using an updated opensource HRV analysis package, the PhysioNet Cardiovascular Signal Toolbox [36]. We extracted the pNNx measures, which represent the percentage of successive normal-to-normal (NN) intervals that differ by more than a threshold of x milliseconds, where x ranged from 5 to 100 ms in increments of 5 ms.

### A.9 Detection of Spindles and Arousals

Sleep spindles and arousals were automatically identified from overnight EEG recordings and sleep-stage annotations.

#### Sleep spindles

Spindle detection was performed using the open-source YASA (Yet Another Spindle Algorithm) Python library [35]. The algorithm applies a multithreshold detection strategy to sigma-band (12–15 Hz) activity from the Fpz EEG channel, extracted from a broadband-filtered (1–30 Hz) signal. Candidate spindles are defined as oscillations lasting 0.5–2 s, separated by at least 500 ms, and exceeding thresholds of relative power (0.2), correlation (0.65), and RMS amplitude (1.5). Detected events were characterized by amplitude (*µ*V), duration (ms), central frequency (Hz), and density (number per minute of N2 sleep).

#### Arousals

Arousal events were estimated from 30-second sleep-stage data based on transitions consistent with the general AASM definition of cortical arousals [34]. Specifically, transitions from N2/N3 to wakefulness, N3 → N1 → N2, and REM → Wake/N1 were classified as arousal events. This transition-based method provides an approximate estimate of AASM-defined arousals using standard hypnogram information. For each recording, total and stage-specific arousals were counted, and arousal indices were expressed as events per hour of total sleep, N2, and N3.

**Table A1:** Participant demographic characteristics (N = 28)

| Characteristic | N (%) | Mean ± SD |
| --- | --- | --- |
| **Age (years)** | | 27.1 ± 4.8 |
| Range | 19 to 37 |  |
| **Sex** |  |  |
| Female | 16 (57.1) |  |
| Male | 12 (42.9) |  |
| **Body Mass Index (kg/m<sup>2</sup>)** | | 25.1 ± 2.9 |
| Range | 20.7 to 30.8 |  |
| **Ethnicity** |  |  |
| Hispanic or Latino | 9 (32.1) |  |
| Not Hispanic or Latino | 19 (67.9) |  |
| **Race** |  |  |
| White | 11 (39.3) |  |
| Asian | 7 (25.0) |  |
| Black or African American | 5 (17.9) |  |
| More than one race | 4 (14.3) |  |
| Unknown or Not Reported | 1 (3.6) |  |
| **Education** |  |  |
| College 4 years or more | 24 (85.7) |  |
| College 1 to 3 years | 4 (14.3) |  |
| **Marital Status** |  |  |
| Single | 23 (82.1) |  |
| Married | 5 (17.9) |  |
| **Employment Status** |  |  |
| Employed | 25 (89.3) |  |
| Unemployed | 3 (10.7) |  |
| **Work Commitment** |  |  |
| Full-time | 12 (42.9) |  |
| Part-time | 9 (32.1) |  |
| None | 7 (25.0) |  |
| **School Commitment** |  |  |
| Full-time | 12 (42.9) |  |
| Part-time | 2 (7.1) |  |
| None | 14 (50.0) |  |

**Figure A1:**
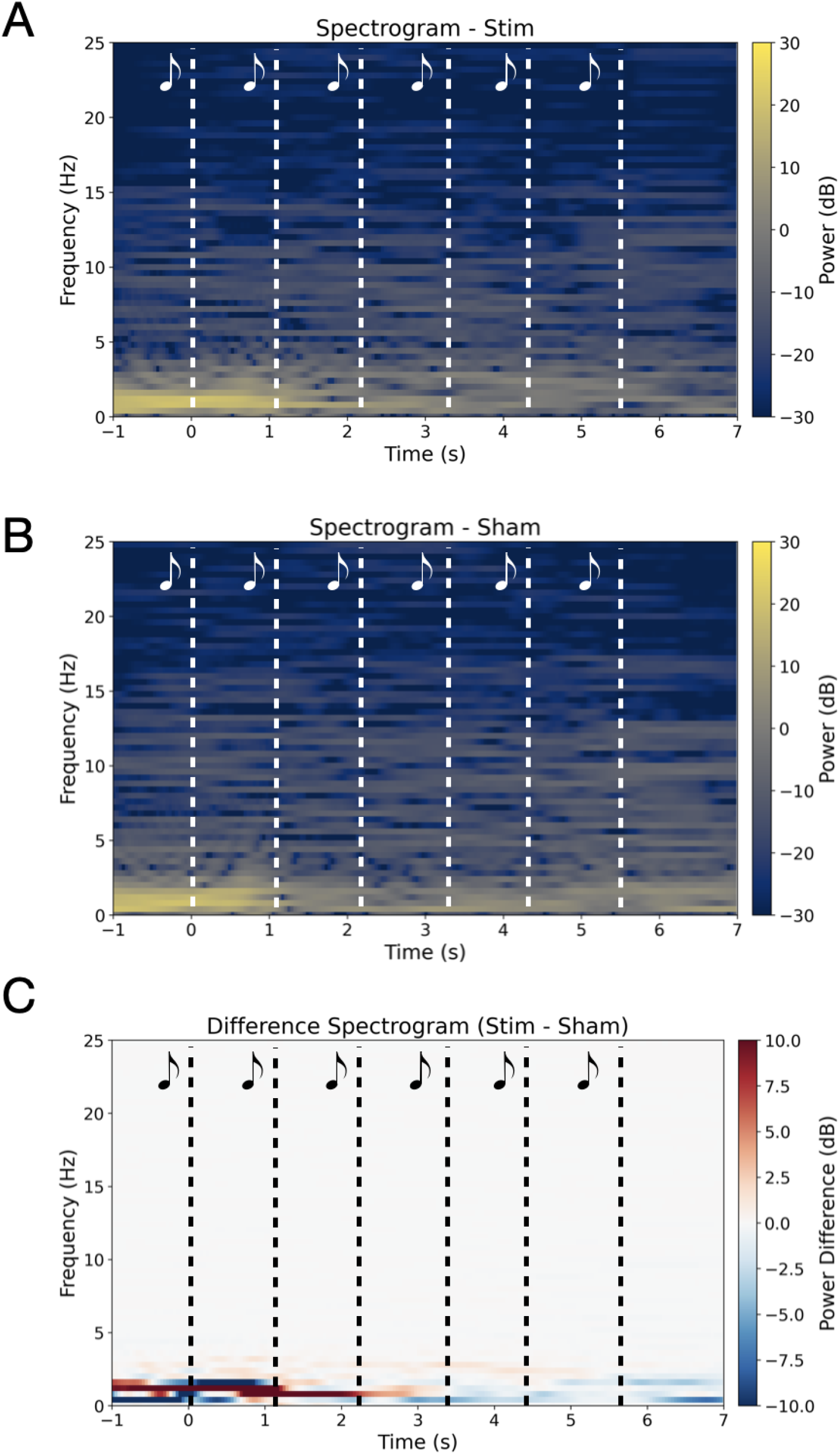
Spectrogram comparison of Stim vs. Sham. (A) Stim condition shows enhanced low-frequency power (<5 Hz) during tone delivery; vertical dashed lines mark tone times, with the strongest increases in the delta band. (B) Sham condition shows no comparable power changes using the same timing markers. (C) Difference spectrogram (Stim − Sham) highlights delta-band (0.5–4 Hz) enhancement shortly after tone onset, indicating modulation of brain activity by auditory stimulation.

**Figure A2:**
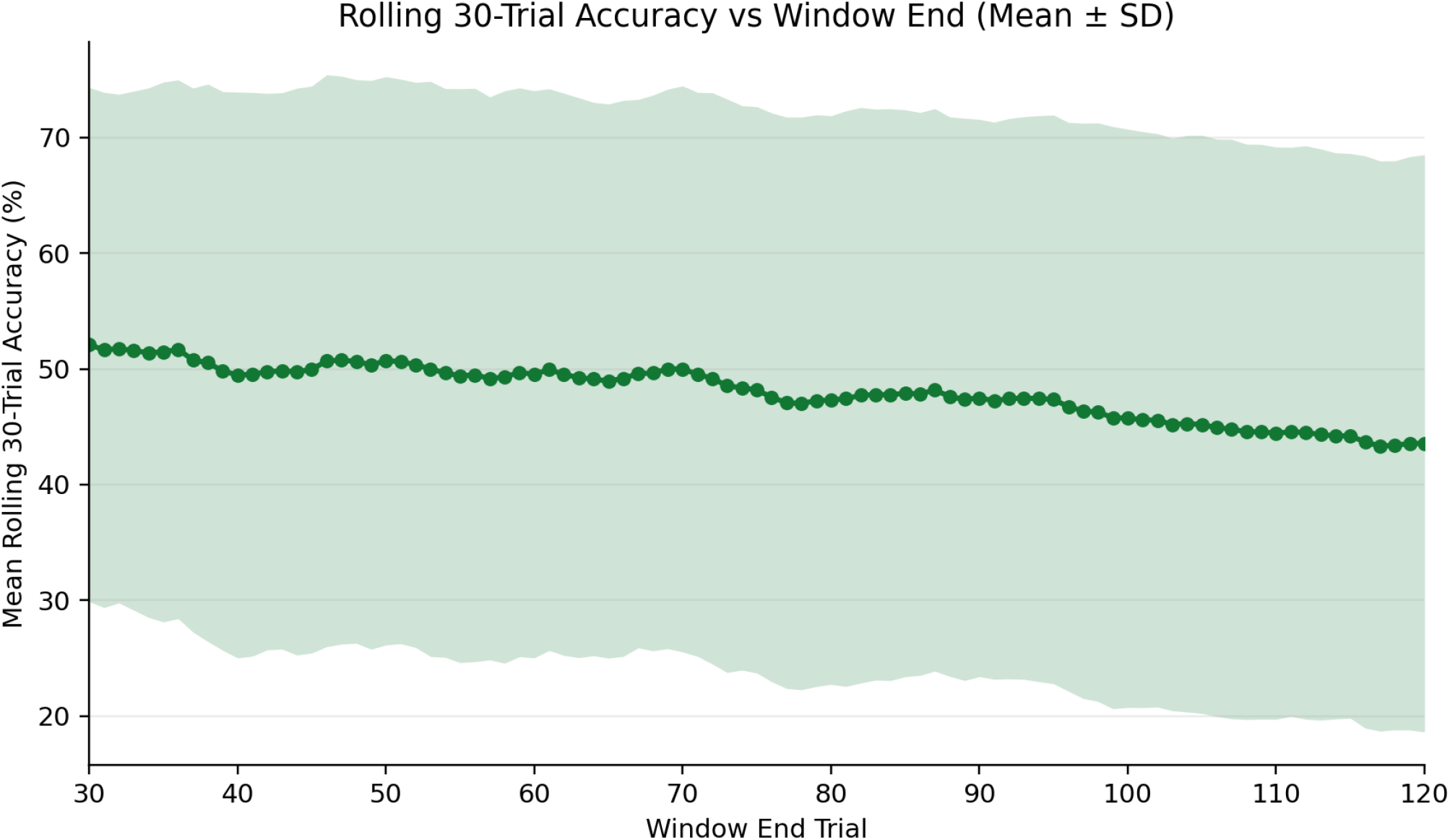
Rolling 30-trial accuracy (mean *±* SD). Points denote windows ending at the indicated trial (1-30 to 91-120).

**Figure A3:**
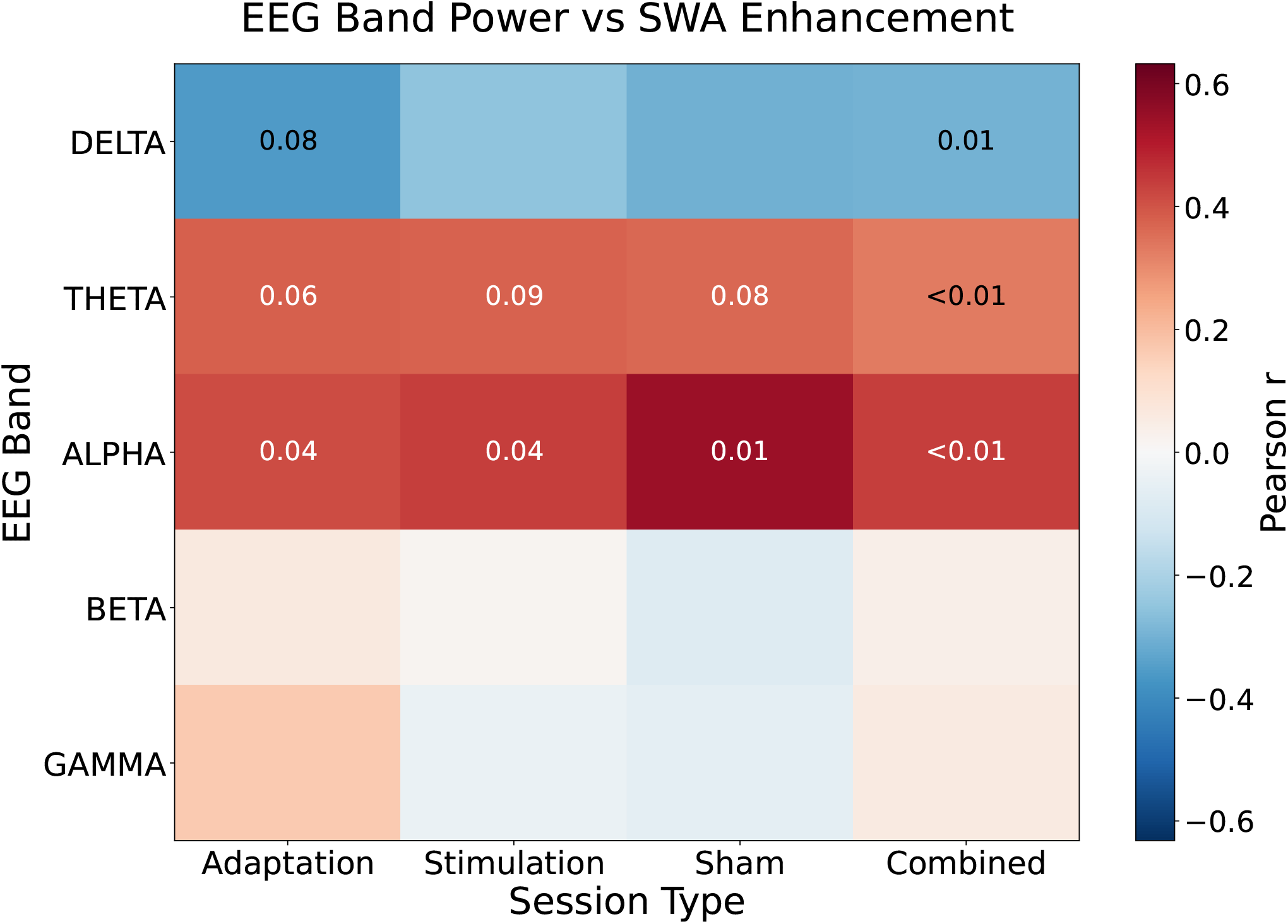
EEG bandpower and SWA Enhancement. Heatmaps showing Pearson correlation coefficients between normalized pre-sleep EEG spectral power in different frequency bands and slow wave activity (SWA) improvement at 1, 4, 7, and 10 hours post-sleep for adaptation, stimulation, sham, and combined sessions. Color intensity indicates correlation strength (red = positive correlation, blue = negative correlation). Only correlations with p ≤ 0.1 are annotated with their p-values.

**Figure A4:**
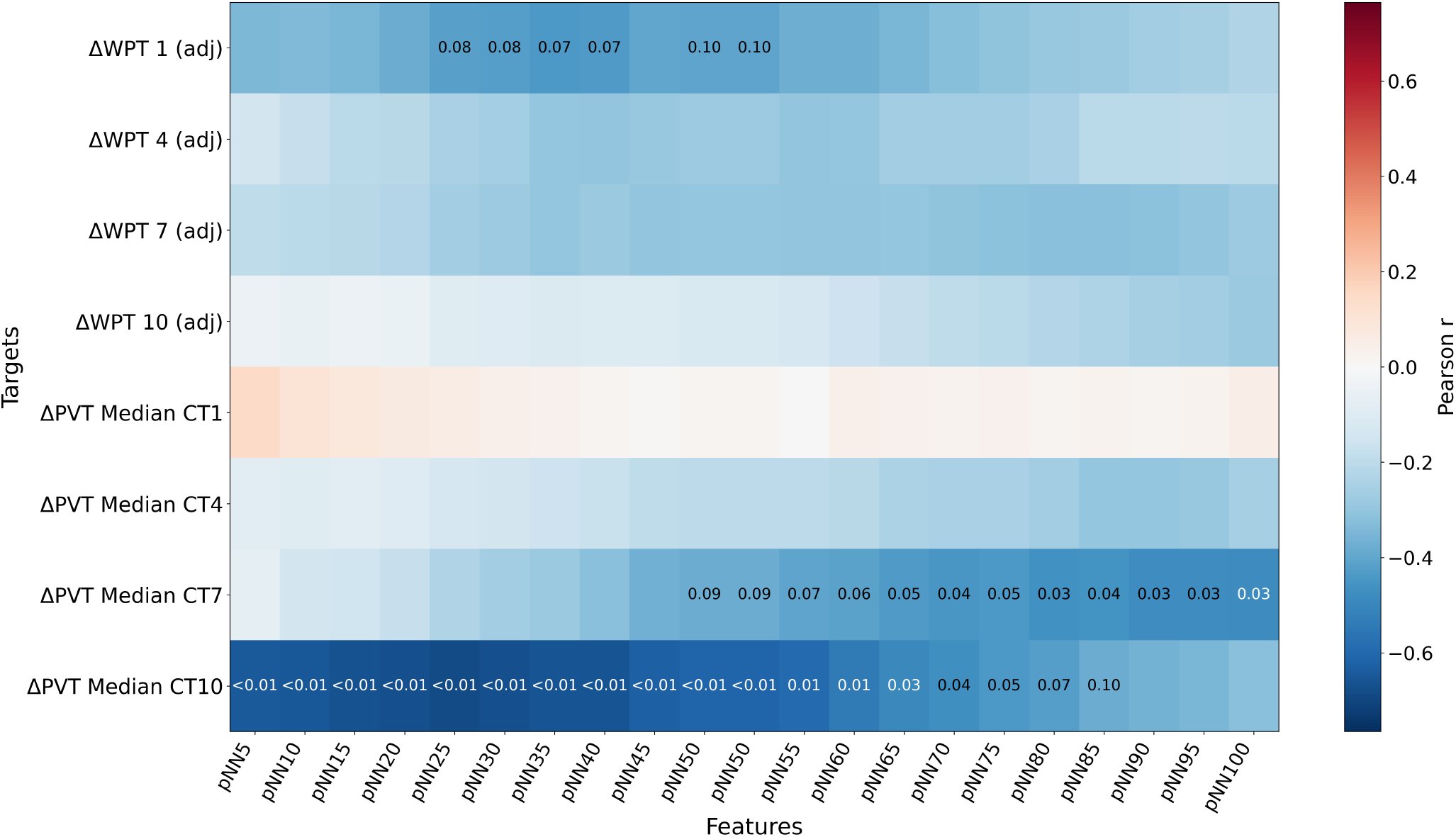
Correlation between pre-sleep heart rate variability metrics and cognitive performance changes. Heatmap showing Pearson correlation coefficients between various HRV metrics (pNNx measures, where x represents different interval thresholds in milliseconds) collected before sleep and changes in word pair task (WPT) and psychomotor vigilance test (PVT) performance at circadian times 1, 4, 7, and 10 hours after wake time. Color intensity indicates correlation strength (blue = negative correlation, red = positive correlation). Only correlations with p ≤ 0.1 are annotated with their correlation coefficient values. pNNx represents the percentage of successive normal-to-normal (NN) intervals that differ by more than x milliseconds, a measure of parasympathetic nervous system activity. ΔWPT and ΔPVT indicate change scores (improvement) from baseline measured before sleep (adj = adjusted).

**Figure A5:**
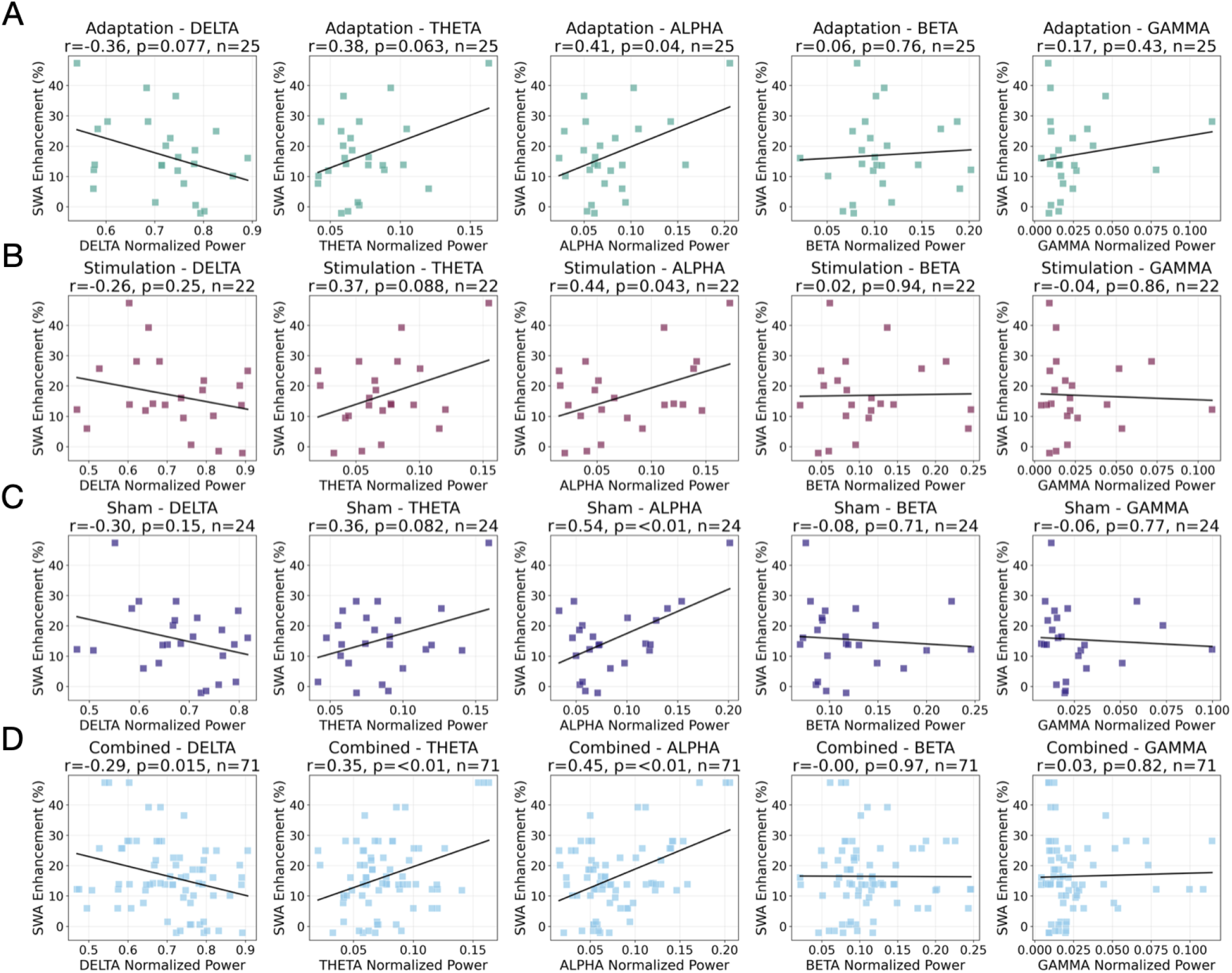
Correlations between pre-sleep EEG power and SWA enhancement across sessions. Scatter plots show associations between normalized pre-sleep EEG band power and slow-wave activity enhancement for (A) Adaptation, (B) Stimulation, (C) Sham, and (D) all sessions combined. Each point represents a participant-night; lines denote linear fits. Correlation coefficients (*r*) and *p*-values are shown in panel titles.

**Figure A6:**
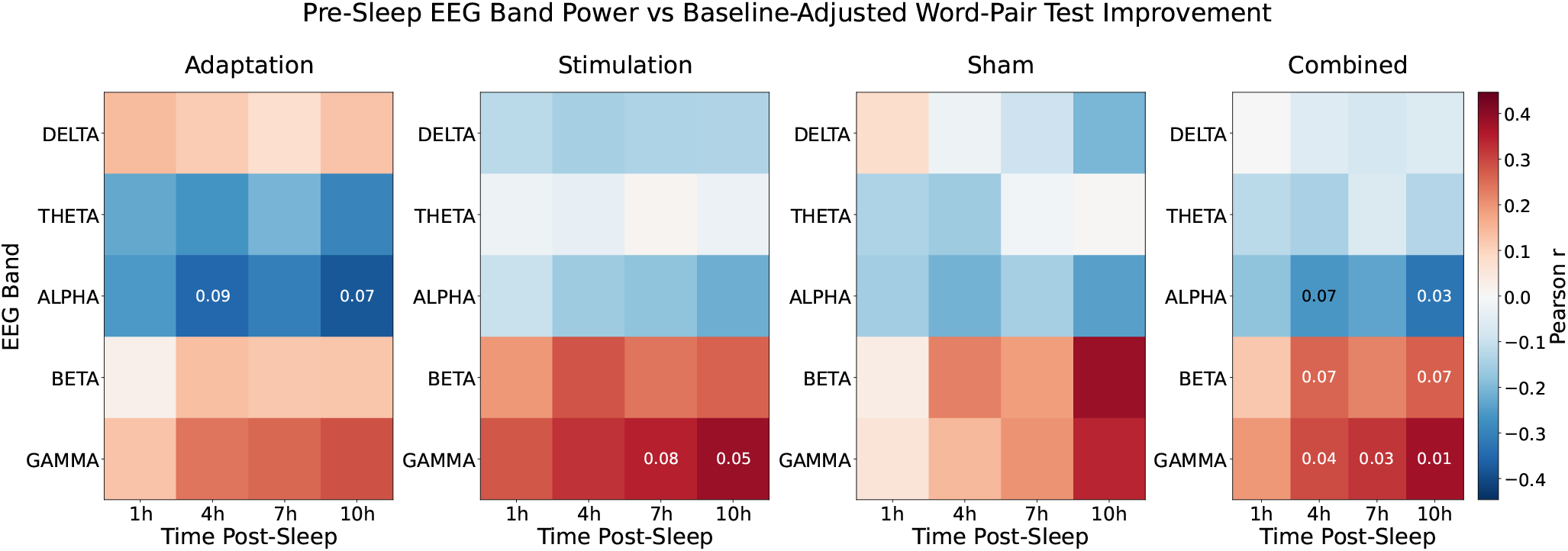
Correlation between pre-sleep EEG band power and baseline-adjusted word-pair test improvement across post-sleep time points. Heatmaps showing Pearson correlation coefficients between normalized pre-sleep EEG spectral power in different frequency bands and recall task improvement at 1, 4, 7, and 10 hours post-sleep for adaptation, stimulation, sham, and combined sessions. Color intensity indicates correlation strength (red = positive correlation, blue = negative correlation). Only correlations with p ≤ 0.1 are annotated with their p-values.

## References

[1] Navya Baranwal, K Yu Phoebe, and Noah S Siegel. Sleep physiology, pathophysiology, and sleep hygiene. Progress in cardiovascular diseases, 77: 59–69, 2023.

[2] Til O Bergmann and Jan Born. Phase-amplitude coupling: a general mechanism for memory processing and synaptic plasticity? Neuron, 97(1): 10–13, 2018.

[3] Luciana Besedovsky, Tanja Lange, and Monika Haack. The sleep-immune crosstalk in health and disease. Physiological reviews, 2019.

[4] Siddharth Biswal, Joshua Kulas, Haoqi Sun, Balaji Goparaju, M Brandon Westover, Matt T Bianchi, and Jimeng Sun. Sleepnet: automated sleep staging system via deep learning. arXiv preprint arXiv:1707.08262, 2017.

[5] Gyorgy Buzsaki and Andreas Draguhn. Neuronal oscillations in cortical networks. science, 304(5679): 1926–1929, 2004.

[6] Roy Cox, Ilia Korjoukov, M. de Boer, and Lucia M. Talamini. Sound asleep: Processing and retention of slow oscillation phase-targeted stimuli. Plos One, 2014.

[7] Ronald E Dahl. The impact of inadequate sleep on children’s daytime cognitive function. In Seminars in pediatric neurology, volume 3, pages 44–50. Elsevier, 1996.

[8] Elizabeth E Devore, Francine Grodstein, Jeanne F Duffy, Meir J Stampfer, Charles A Czeisler, and Eva S Schernhammer. Sleep duration in midlife and later life in relation to cognition. Journal of the American Geriatrics Society, 62(6): 1073–1081, 2014.

[9] Charmaine Diep, Suzanne Ftouni, Jessica E Manousakis, Christian L Nicholas, Sean PA Drummond, and Clare Anderson. Acoustic slow wave sleep enhancement via a novel, automated device improves executive function in middle-aged men. Sleep, 43(1):zsz197, 2020.

[10] M Dresler, VI Spoormaker, P Beitinger, M Czisch, M Kimura, A Steiger, and F Holsboer. Neurosciencedriven discovery and development of sleep therapeutics. Pharmacology & therapeutics, 141(3): 300–334, 2014.

[11] Marcos G Frank and H Craig Heller. The function (s) of sleep. Sleep-wake neurobiology and pharmacology, pages 3–34, 2019.

[12] Lisa Genzel, Victor I. Spoormaker, Boris N. Konrad, and Martin Dresler. The role of rapid eye movement sleep for amygdala-related memory processing. Neurobiology of Learning and Memory, 2015.

[13] Theresa E Gildner, Aarón Salinas-Rodríguez, Betty Manrique-Espinoza, Karla Moreno-Tamayo, and Paul Kowal. Does poor sleep impair cognition during aging? longitudinal associations between changes in sleep duration and cognitive performance among older mexican adults. Archives of gerontology and geriatrics, 83: 161–168, 2019.

[14] Shojiro Inoué, Kazuki Honda, and Yasuo Komoda. Sleep as neuronal detoxification and restitution. Behavioural brain research, 69(1–2): 91–96, 1995.

[15] Anna Ivanenko and Kyle Johnson. Sleep disturbances in children with psychiatric disorders. In Seminars in Pediatric Neurology, volume 15, pages 70–78. Elsevier, 2008.

[16] Jens G Klinzing, Niels Niethard, and Jan Born. Mechanisms of systems memory consolidation during sleep. Nature neuroscience, 22(10): 1598–1610, 2019.

[17] Minchao Li, Nan Wang, and Matthew E Dupre. Association between the self-reported duration and quality of sleep and cognitive function among middle-aged and older adults in china. Journal of Affective Disorders, 304: 20–27, 2022.

[18] Lisa Marshall, Halla Helgadóttir, Matthias Mölle, and Jan Born. Boosting slow oscillations during sleep potentiates memory. Nature, 444(7119):610–613, 2006.

[19] Seiko Miyata, Akiko Noda, Norio Ozaki, Yuki Hara, Makoto Minoshima, Kunihiro Iwamoto, Masahiro Takahashi, Tetsuya Iidaka, and Yasuo Koike. Insufficient sleep impairs driving performance and cognitive function. Neuroscience letters, 469(2): 229–233, 2010.

[20] Miguel Navarrete, Jules Schneider, Hong-Viet V. Ngo, Mario Valderrama, Alexander J. Casson, and Penelope A. Lewis. Examining the optimal timing for closed-loop auditory stimulation of slow-wave sleep in young and older adults. Sleep, 2019.

[21] Garrett T Neske. The slow oscillation in cortical and thalamic networks: mechanisms and functions. Frontiers in neural circuits, 9:88, 2016.

[22] Hong Viet Ngo, Thomas Martinetz, Jan Born, and Matthias Mölle. Auditory closed-loop stimulation of the sleep slow oscillation enhances memory. Neuron, 2013.

[23] Hong-Viet V Ngo, Thomas Martinetz, Jan Born, and Matthias Mölle. Auditory closed-loop stimulation of the sleep slow oscillation enhances memory. Neuron, 78(3): 545–553, 2013.

[24] Ju Lynn Ong, June C Lo, Nicholas IYN Chee, Giovanni Santostasi, Ken A Paller, Phyllis C Zee, and Michael WL Chee. Effects of phase-locked acoustic stimulation during a nap on eeg spectra and declarative memory consolidation. Sleep Medicine, 20:88–97, 2016.

[25] Jessica E Owen and Sigrid C Veasey. Impact of sleep disturbances on neurodegeneration: Insight from studies in animal models. Neurobiology of disease, 139:104820, 2020.

[26] Nelly A Papalambros, Sandra Weintraub, Tammy Chen, Daniela Grimaldi, Giovanni Santostasi, Ken A Paller, Phyllis C Zee, and Roneil G Malkani. Acoustic enhancement of sleep slow oscillations in mild cognitive impairment. Annals of clinical and translational neurology, 6(7): 1191–1201, 2019.

[27] B. Rasch and J. Born. About sleep’s role in memory. Physiological Reviews, 93: 681–766, 2013.

[28] Björn Rasch and Jan Born. About sleep’s role in memory. Physiological Reviews, 2013.

[29] Frank Raven, Eddy A Van der Zee, Peter Meerlo, and Robbert Havekes. The role of sleep in regulating structural plasticity and synaptic strength: implications for memory and cognitive function. Sleep medicine reviews, 39: 3–11, 2018.

[30] Giovanni Santostasi, Roneil G. Malkani, Brady A. Riedner, Michele Bellesi, Giulio Tononi, Ken A. Paller, and Phyllis C. Zee. Phase-locked loop for precisely timed acoustic stimulation during sleep. Journal of Neuroscience Methods, 2016.

[31] Mihkel Stamm, Jaan Aru, Renate Rutiku, and Talis Bachmann. Occipital long-interval paired pulse tms leads to slow wave components in nrem sleep. Consciousness and Cognition, 35: 78–87, 2015.

[32] Akara Supratak, Hao Dong, Chao Wu, and Yike Guo. Deepsleepnet: A model for automatic sleep stage scoring based on raw single-channel eeg. IEEE transactions on neural systems and rehabilitation engineering, 25(11): 1998–2008, 2017.

[33] Akara Supratak and Yike Guo. Tinysleepnet: An efficient deep learning model for sleep stage scoring based on raw single-channel eeg. In 2020 42nd Annual International Conference of the IEEE Engineering in Medicine & Biology Society (EMBC), pages 641–644. IEEE, 2020.

[34] Matthew M. Troester, Stuart F. Quan, Richard B. Berry, et al. The AASM Manual for the Scoring of Sleep and Associated Events: Rules, Terminology, and Technical Specifications. American Academy of Sleep Medicine, Darien, IL, version 3 edition, 2023. for the American Academy of Sleep Medicine.

[35] Raphael Vallat and Matthew P Walker. An opensource, high-performance tool for automated sleep staging. elife, 10:e70092, 2021.

[36] Adriana N Vest, Giulia Da Poian, Qiao Li, Chengyu Liu, Shamim Nemati, Amit J Shah, and Gari D Clifford. An open source benchmarked toolbox for cardiovascular waveform and interval analysis. Physiological measurement, 39(10): 105004, 2018.

[37] Chanung Wang and David M Holtzman. Bidirectional relationship between sleep and alzheimer’s disease: role of amyloid, tau, and other factors. Neuropsychopharmacology, 45(1): 104–120, 2020.

[38] Arne Weigenand, Matthias Mölle, Friederike Werner, Thomas Martinetz, and Lisa Marshall. Timing matters: open-loop stimulation does not improve overnight consolidation of word pairs in humans. European Journal of Neuroscience, 44(6): 2357–2368, 2016.

[39] Carmen E Westerberg, Susan M Florczak, Sandra Weintraub, M-Marsel Mesulam, Lisa Marshall, Phyllis C Zee, and Ken A Paller. Memory improvement via slow-oscillatory stimulation during sleep in older adults. Neurobiology of aging, 36(9): 2577–2586, 2015.

[40] Lulu Xie, Hyunmo Kang, Qiwu Xu, Michael Chen, Yonghong Liao, Meenakshisundaram Thiyagarajan, John O’Donnell, Daniel J. Christensen, Charles Nicholson, Jeffrey J. Iliff, Takahiro Takano, and Rashid Deane. Sleep drives metabolite clearance from the adult brain. Science, 2013.

